# The cellular immune response to COVID-19 deciphered by single cell multi-omics across three UK centres

**DOI:** 10.1101/2021.01.13.21249725

**Authors:** Emily Stephenson, Gary Reynolds, Rachel A Botting, Fernando J Calero-Nieto, Michael Morgan, Zewen Kelvin Tuong, Karsten Bach, Waradon Sungnak, Kaylee B Worlock, Masahiro Yoshida, Natsuhiko Kumasaka, Katarzyna Kania, Justin Engelbert, Bayanne Olabi, Jarmila Stremenova Spegarova, Nicola K Wilson, Nicole Mende, Laura Jardine, Louis CS Gardner, Issac Goh, Dave Horsfall, Jim McGrath, Simone Webb, Michael W. Mather, Rik GH Lindeboom, Emma Dann, Ni Huang, Krzysztof Polanski, Elena Prigmore, Florian Gothe, Jonathan Scott, Rebecca P Payne, Kenneth F Baker, Aidan T Hanrath, Ina CD Schim van der Loeff, Andrew S Barr, Amada Sanchez-Gonzalez, Laura Bergamaschi, Federica Mescia, Josephine L Barnes, Eliz Kilich, Angus de Wilton, Anita Saigal, Aarash Saleh, Sam M Janes, Claire M Smith, Nusayhah Gopee, Caroline Wilson, Paul Coupland, Jonathan M Coxhead, Vladimir Y Kiselev, Stijn van Dongen, Jaume Bacardit, Hamish W King, Cambridge Institute of Therapeutic Immunology and Infectious Disease-National Institute of Health Research (CITIID-NIHR) COVID BioResource Collaboration, Anthony J Rostron, A John Simpson, Sophie Hambleton, Elisa Laurenti, Paul A Lyons, Kerstin B Meyer, Marko Z Nikolic, Christopher JA Duncan, Ken Smith, Sarah A Teichmann, Menna R Clatworthy, John C Marioni, Berthold Gottgens, Muzlifah Haniffa

## Abstract

The COVID-19 pandemic, caused by SARS coronavirus 2 (SARS-CoV-2), has resulted in excess morbidity and mortality as well as economic decline. To characterise the systemic host immune response to SARS-CoV-2, we performed single-cell RNA-sequencing coupled with analysis of cell surface proteins, providing molecular profiling of over 800,000 peripheral blood mononuclear cells from a cohort of 130 patients with COVID-19. Our cohort, from three UK centres, spans the spectrum of clinical presentations and disease severities ranging from asymptomatic to critical. Three control groups were included: healthy volunteers, patients suffering from a non-COVID-19 severe respiratory illness and healthy individuals administered with intravenous lipopolysaccharide to model an acute inflammatory response. Full single cell transcriptomes coupled with quantification of 188 cell surface proteins, and T and B lymphocyte antigen receptor repertoires have provided several insights into COVID-19: 1. a new non-classical monocyte state that sequesters platelets and replenishes the alveolar macrophage pool; 2. platelet activation accompanied by early priming towards megakaryopoiesis in immature haematopoietic stem/progenitor cells and expansion of megakaryocyte-primed progenitors; 3. increased clonally expanded CD8^+^ effector:effector memory T cells, and proliferating CD4^+^ and CD8^+^ T cells in patients with more severe disease; and 4. relative increase of IgA plasmablasts in asymptomatic stages that switches to expansion of IgG plasmablasts and plasma cells, accompanied with higher incidence of BCR sharing, as disease severity increases. All data and analysis results are available for interrogation and data mining through an intuitive web portal. Together, these data detail the cellular processes present in peripheral blood during an acute immune response to COVID-19, and serve as a template for multi-omic single cell data integration across multiple centers to rapidly build powerful resources to help combat diseases such as COVID-19.

## Introduction

The outbreak of coronavirus disease 2019 (COVID-19) was declared a global pandemic on 11 March 2020^1^, and as of 12 January 2021 has led to over 91 million infections and 1.9 million deaths worldwide^2^. Common symptoms, which are often mild and transient, include cough, fever and loss of taste and/or smell^3^. In a small proportion of those infected, symptoms can worsen and lead to hospitalisation, with the elderly and those with comorbidities being most at risk^4^. In critical cases, patients may develop acute respiratory distress syndrome (ARDS) necessitating intensive care therapy, including endotracheal intubation and mechanical ventilation^5^. Clinical trials of vaccines and therapeutics have been performed at an unprecedented pace^6^, leading to the emergency authorisation of several vaccine candidates for susceptible populations in December 2020^7^. Treatment strategies under investigation include medication with anti-viral, anti-inflammatory and immunomodulatory properties^8^.

The aetiologic agent of COVID-19 is a novel highly-infectious pathogenic coronavirus, severe acute respiratory syndrome coronavirus 2 (SARS-CoV-2). This enveloped, positive-sense single-stranded RNA betacoronavirus utilises the cell surface receptor angiotensin-converting enzyme 2 (ACE2) to enter host cells^9^. ACE2 is expressed in various barrier tissues, including nasal epithelium, conjunctival epithelium and intestines, as well as internal organs, including alveoli of the lung, heart, brain, kidney and the uterine-placental interface^10^. Neuropilin (NRP1), a cell surface receptor expressed in respiratory and olfactory epithelium, can also facilitate SARS-CoV-2 cellular entry^11^. Patients with COVID-19 infection often have lymphopenia in association with high neutrophil and platelet counts, parameters which may give prognostic indication^12^.

Several studies have highlighted a complex network of peripheral blood immune responses in COVID-19 infection, with the role of T cells during infection being an area of particular focus^13,14^. A reduction of absolute numbers of T cells linked with disease severity has been reported, as well as a decrease in IFN-γ production by lymphocytes^15^. However, a significant expansion of highly cytotoxic effector T cell subsets has also been found in patients with moderate disease^16^. Additionally, higher expression of exhaustion markers PD-1 and Tim-3 on CD8^+^ T cells have been described in patients receiving Intensive Care Unit (ICU) therapy^17^.

The response of myeloid cells and B cells have been less well explored in COVID-19. Emergency myelopoiesis, driven by inflammation, is thought to arise as a way to prevent tissue damage^18,19^. In severe cases, dysregulation of myelopoiesis coupled with abnormal monocyte activation can occur^18^, but the underlying mechanisms remain to be explored. Extrafollicular B cell activation is present in critically ill patients but despite the high levels of SARS-CoV-2 specific antibodies and antibody secreting cells, many of these patients do not recover from the disease^14^. Neutralising antibodies are protective against infection and potentially confer immunity to reinfection, as adoptive transfer of anti-SARS-CoV-2 monoclonal antibodies into naive animals were shown to reduce virus replication and disease development^20^. Vaccine-induced neutralising antibody titers have been correlated with protection in nonhuman primates^21^, and recovered patients display robust antibody responses correlated with neutralisation of authentic virus for at least several months^22^.

In this study we combined single cell resolution analysis of transcriptomes, cell surface proteins and lymphocyte antigen receptor repertoires to characterise the cellular immune response in peripheral blood to COVID-19 across a range of disease severities, integrating results across three UK medical centres.

## Results

### Altered transcriptomic and surface protein profiles related to severity of COVID-19 infection

To delineate the immune response to COVID-19 infection, we collected venous blood samples from patients with asymptomatic, mild, moderate, severe and critical^23^ COVID-19 infection across three UK centres in Newcastle, Cambridge and London. Controls included healthy volunteers, patients with a non-COVID-19 severe respiratory illness, and healthy volunteers administered with intravenous lipopolysaccharide (IV-LPS) as a surrogate for an acute systemic bacterial inflammatory response (**Fig. 1A, Supplementary Table 1**). We generated single cell transcriptome data from peripheral blood mononuclear cells (PBMCs) of all individuals as well as a census of cell surface proteins using a panel of 192 antibody derived tags (ADT) (**Fig. 1A, Supplementary Table 2**). In total, following demultiplexing and doublet removal, we sequenced 1,141,860 cells from 143 samples with 850,100 cells passing quality control (min of 200 genes and <10% mitochondrial reads/cell) **(Extended Data 1A)**. The full scRNA-seq dataset was integrated using Harmony^24^ (**Fig. 1B**). There was good mixing of cells by the kBET statistic calculated for each cluster across sample IDs (rejection rate improved from 0.62 to 0.36 following integration, p<2.1×10^−8^ by Wilcoxon paired signed rank test (**Extended Data 1B-C**)).

**Figure 1:**
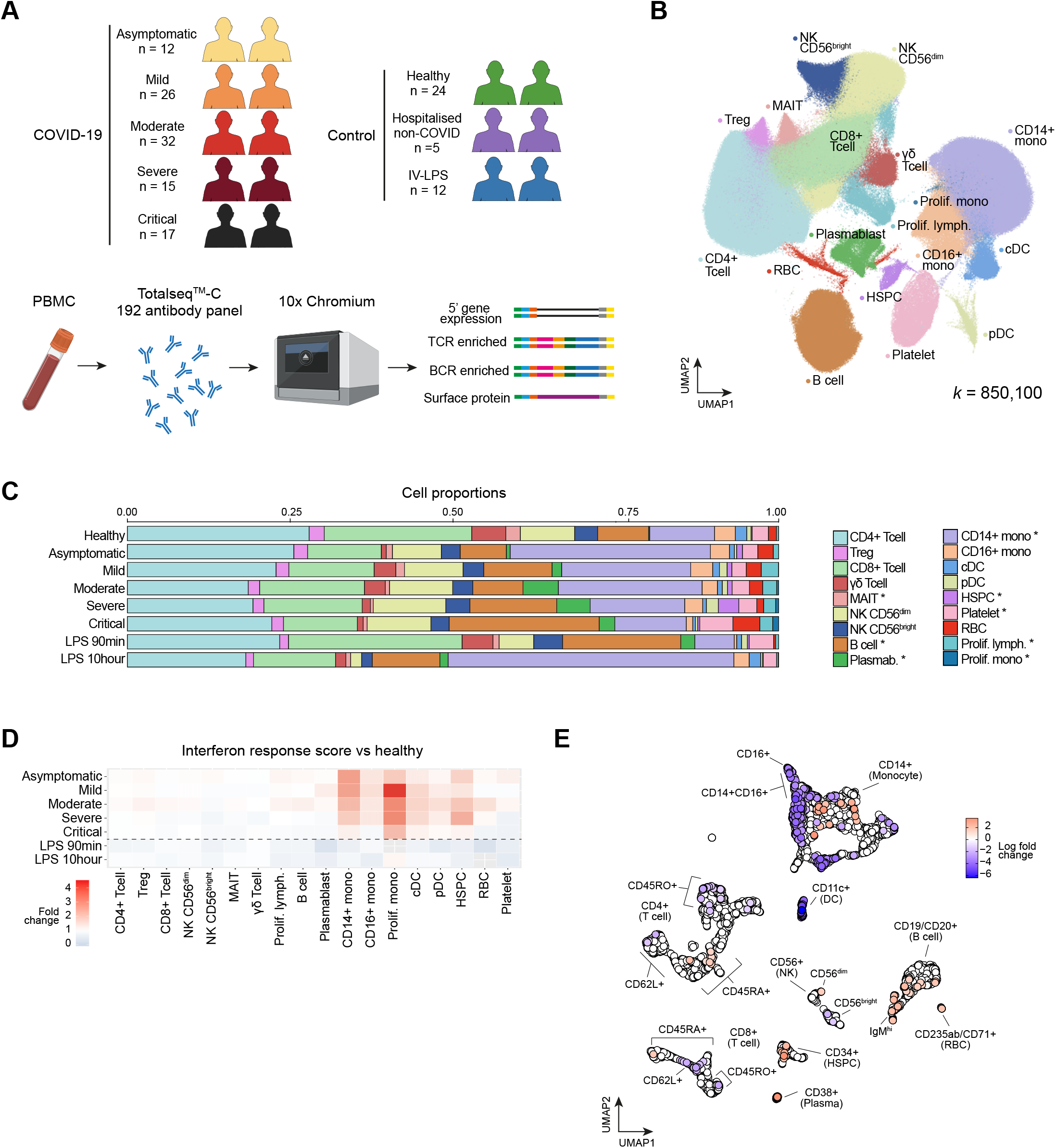
Single cell multi-omic analysis of COVID-19 patients’ PBMC. **A.** Overview of the participants included and the samples and data collected. IV-LPS, intravenous lipopolysaccharide; PBMC, peripheral blood mononuclear cells. **B**. UMAP visualisation of all 850,100 cells sequenced. Leiden clusters based on 5’ gene expression shown and coloured by cell type. DC, dendritic cell; HSPC, haematopoietic stem and progenitor cell; lymph, lymphocyte; MAIT, mucosal-associated invariant T cell; mono, monocyte; pDC, plasmacytoid dendritic cell; Prolif., proliferating; RBC, red blood cell; NK, natural killer cell. **C**. Bar plot of the proportion of cell types shown in **B**. separated by condition and COVID-19 severity status. Hypothesis testing was performed using quasi-likelihood F-test comparing healthy controls to cases for linear trends across disease severity groups (healthy > asymptomatic > mild > moderate > severe > critical). Differentially abundant cell types were determined using a 10% false discovery rate (FDR) and marked (*). **D**. Enrichment of interferon response of each cell state separated by severity. IFN response was calculated using a published gene list (GO: 0034340) **E**. UMAP computed using batch-corrected mean staining intensities of 188 antibodies for 4241 hyperspheres. Each hypersphere represents an area in the 188-dimensional space and is colored by significant (spatial FDR < 0.05) severity associated changes in abundance of cells within that space.

Following Leiden clustering, cells were manually annotated based on the RNA expression of known marker genes. RNA-based annotation was supported by surface protein expression of markers commonly employed in flow cytometry to discriminate PBMC subpopulations (**Extended Data 1D**). We defined 18 cell subsets across the datasets (**Fig. 1B**), with an additional 27 cell states identified following sub-clustering (**Fig. 1B, 2A, 3A-B, 4A-B**). Our annotation was further validated using the Azimuth annotation tool for PBMC where more than 50% of the cells were mapped and matched to a unique cluster in 32/33 of the clusters defined in the Azimuth PBMC dataset (proliferating CD8 T cells mapped across two clusters). Clusters unique to our data include proliferating monocytes, ILC subpopulations and isotype-specific plasma cells. (**Extended Data 1E**). Our complete COVID-19 peripheral blood multi-omic data is available through the web-portal at https://covid19cellatlas.org.

We assessed how cell populations varied with severity of COVID-19 and observed a relative expansion of proliferating lymphocytes, proliferating monocytes, platelets, and mobilized haematopoietic stem/progenitor cells (HSPCs) with worsening disease. In the B cell compartment, there was an expansion of plasmablasts in COVID-19 and an increase in B cells in severe and critical disease. In contrast to these expansions, there were reductions in MAIT cells with disease severity (**Fig. 1C, Extended Data 2A**). These changes were in keeping with the trends observed in clinical blood lymphocyte, monocyte and platelet counts of COVID-19 patients (**Extended Data 2B, Supplementary Table 3**). To assess the broader impacts of patient characteristics and clinical metadata on the altered proportion of cell type/states, we used a Poisson linear mixed model (see Methods and **Supplementary Note 1**) which predicted the COVID-19 swab result (BF corrected LR *P*=2.3×10^−4^; see Methods), disease severity at blood sampling (BF corrected LR *P*=3.5×10^−7^), and centre (contributed by increased RBC and reduced monocytes in the Cambridge patient cohort; BF corrected LR *P*=3.5×10^−142^) as the main contributing factors to cell population frequency among 7 different clinical/technical factors (**Extended Data 2C**). Further, we found that PBMC composition varied depending on symptom duration, with increased relative frequency of pDCs, NK cells, CD14+ and CD16+ monocytes (FDR 10%) and decreased relative frequency of B cells, Tregs, RBCs, platelets and CD4 T cells with a longer symptomatic interval before sampling (**Extended Data 2E**). Critically ill patients were sampled at later time points from onset of symptoms than mild-moderate-severe patients, consistent with the protracted course of infection in critical disease (**Extended Data 2D**). However, concordant changes according to symptom duration were still found when excluding critical patients, indicating the additional influence of symptom duration on peripheral immune cell changes in SARS-CoV-2 infection (**Extended Data 2F**).

We observed expression of Type I/III interferon response genes in monocytes, DCs and HSPCs across the spectrum of COVID-19 severity, but not in patients challenged with IV-LPS, in keeping with the importance of type I and III interferons in the innate immune response to viral infection (**Fig. 1D**). Type I/III interferon response-related genes were recently identified as harbouring association signals in a Genome Wide Association Study (GWAS) for COVID-19 susceptibility^25,26^. Of the genes identified in this study, we found *IFNAR2* was both upregulated in COVID-19 compared to healthy in most circulating cell types and highly expressed by plasmablasts, monocytes and DCs (**Extended Data 2G**).

To provide information on the cytokine and chemokine context influencing peripheral immune cells, we performed multiplexed analysis of 45 proteins in serum. Two contrasting cytokine profiles were evident when comparing mild/moderate to severe/critical patients. CCL4, CXCL10, IL7 and IL1A were associated with severe and critical disease, suggesting an augmented drive for monocyte and NK lymphocyte recruitment as well as support for T cell activity/pathology (**Extended Data 2H, Supplementary Table 4**).

To take advantage of the comprehensive protein expression data, we used Cydar^27^ to characterise how the immune landscape changes with disease severity based on surface protein expression. We divided cells into phenotypic hyperspheres based on the expression of 188 proteins. We then quantified the number of cells from each severity group within the hyperspheres, which allowed us to identify 430 hyperspheres that differed significantly in abundance with increasing severity (spatial FDR < 0.05, **Fig. 1E**). Examining the surface protein expression profiles post-hoc showed that differentially abundant hyperspheres were present in all major immune compartments. In particular, we found an increase in populations of B cells (CD19^+^/CD20^+^), plasma cells (CD38^+^) and HSPCs (CD34^+^) as well as a previously reported remodelling of the myeloid compartment^18^ (**Fig. 1E**).

### Mononuclear phagocytes and haematopoietic stem progenitors

Transcriptome and surface proteome analysis of blood mononuclear phagocytes identified known DC subsets (pDC, ASDC, DC1, DC2, DC3) and several monocyte states (**Figs. 2A-B**). Three cell states of CD14^+^ monocytes are present (proliferating, classical CD14^+^ and activated CD83^+^) in addition to two CD14^+^CD16^+^ monocyte cell states (non-classical CD16^+^ and C1QA/B/C^+^) (**Figs. 2A-B)**. Proliferating monocytes and DCs expressing *MKI67* and *TOP2A* are present in increasing frequency with worsening severity of COVID-19 (**Figs. 2A-B**). In contrast, circulating numbers of DC2 and DC3 are reduced. Proliferating monocytes have previously been identified by flow cytometry of COVID-19 patients’ blood^19^. Here, we add that they share an extended protein and RNA expression profile with CD14^+^ monocytes (**Figs. 2A-B)**. Proliferating DCs most closely resemble DC2. *C1QA/B/C*-expressing CD16^+^ monocytes are present at a low frequency relative to whole PBMC in healthy blood, but are expanded in COVID-19 and are the only source of C1 complement components in PBMCs (**Fig. 2B, Extended Data 3A**).

**Figure 2:**
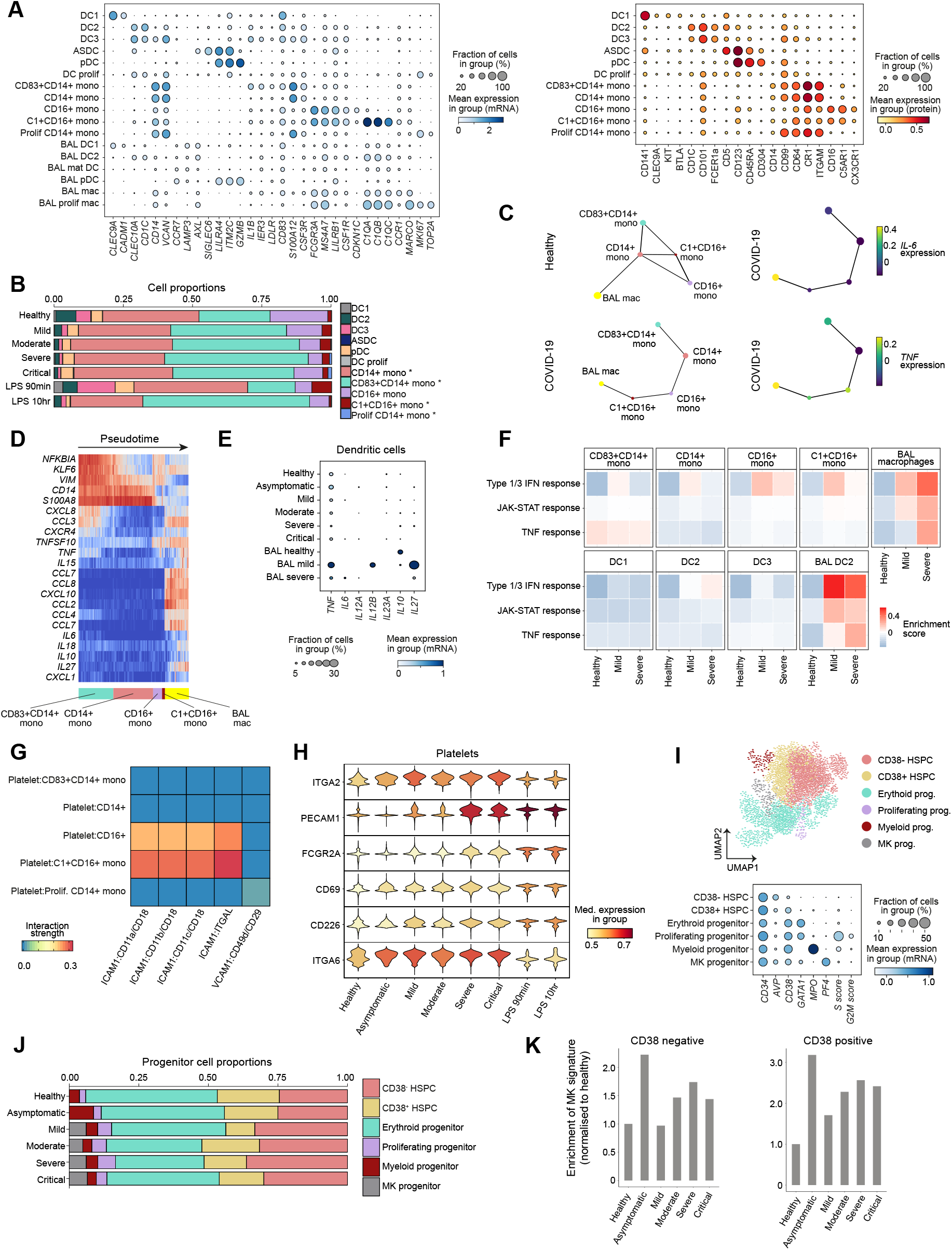
Myeloid and progenitor cells. **A.** Dot plots of gene expression (left; blue) and surface protein (right; red) expression for myeloid populations where the colour is scaled by mean expression and the dot size is proportional to the percent of the population expressing the gene/protein, respectively. **B**. Bar plot of the proportion of myeloid populations separated by condition and severity status from the Ncl and Sanger/UCL site. Hypothesis testing was performed using quasi-likelihood F-test comparing healthy controls to cases. Differentially abundant cell types were determined using a 10% false discovery rate (FDR) and marked (*). **C**. Partition based graph abstraction (PAGA) representing connectivity between clusters defined in **A** for healthy (top left) and COVID-19 (bottom left) monocytes and BAL macrophages. Expression of *IL6* (top right) and *TNF* (bottom right) in each cluster along the predicted path for COVID-19 monocytes. **D**. Expression of differentially expressed cytokines between CD83^+^CD14^+^ monocytes and BAL macrophages shown by cells ordered by pseudotime calculated for COVID-19 monocytes and BAL macrophages from **C. E**. Expression of DC-derived T cell polarising cytokines in peripheral blood DC2 and mature BAL DCs. **F**. Heat map displaying gene set enrichment scores for Type 1/3 IFN-response, TNF-response and JAK-STAT signatures in the myeloid populations found in COVID-19 PBMCs. **G**. Heat map of CellPhoneDB predicted ligand:receptor interactions between platelets and monocyte subsets. **H**. Violin plots showing significantly differentially expressed markers of platelet activation proteins between healthy and COVID-19. **I**. UMAP representation of HSPCs (top) and gene expression markers used to annotate clusters (below). MK, Megakaryocyte **J**. Bar chart of the proportion of progenitors by severity status. MK, Megakaryocyte. **K**. Bar charts displaying enrichment of a megakaryocyte signature found in CD34^-^CD38^-^ (left) and CD34^+^CD38^+^ HSPCs (right), separated by severity. MK, megakaryocyte.

We previously demonstrated the egress of blood DCs and monocytes from blood to alveolar space with rapid acquisition of a lung molecular profile following human inhalational LPS challenge^28^. To better understand the relationship between circulating and lung alveolar mononuclear phagocytes in COVID-19, we compared the transcriptome profile of blood DCs and monocytes with their bronchoalveolar lavage (BAL) counterparts during COVID-19 using recently published data (GSE145926)^29^ (**Extended Data 3B**). As expected, partition-based graph abstraction (PAGA) suggests transcriptional similarity between healthy circulating CD14^+^ monocytes and healthy BAL macrophages, in agreement with recent data demonstrating that BAL macrophages can arise from circulating CD14^+^ monocytes (**Fig. 2C**)^30^. However, there is a surprisingly greater transcriptional similarity between BAL macrophages and the expanded population of circulating C1QA/B/C^+^CD16^+^ monocytes in COVID-19 (**Fig. 2C**). These observations raise the possibility of a differential origin of alveolar macrophages during health and COVID-19. Both BAL macrophages and C1QA/B/C^+^CD16^+^ monocytes express *FCGR3A* and *C1QA/B/C* and are enriched for expression of type I interferon response genes (**Fig. 2A**). Myeloid hyperinflammatory response has been reported to mediate lung and peripheral tissue damage via secretion of inflammatory cytokines such as IL-6 and TNFa in COVID-19. We evaluated the expression of these cytokines and found that they are primarily expressed by tissue rather than blood mononuclear phagocytes (**Fig. 2C**). Genes differentially expressed in CD83^+^ CD14^+^ monocytes and BAL macrophages across pseudotime identified expression of *IL15*, which is produced in response to viral infections to promote NK proliferation, and leukocyte recruiting chemokines including *CCL2, CCL4, CCL7*, and *CCL8* upregulated by BAL macrophages (**Fig. 2D**).

Tissue DCs respond to local inflammation and pathogen challenge by migrating to the draining lymph node to activate naïve T cells. BAL contains a population of mature, migratory DCs that express *CCR7* and *LAMP3* but downregulate DC-specific markers such as *CD1C* and *CLEC9A* (**Extended Data 3B**). These migratory DCs express *IL10* in healthy BAL but express *TNF* and the common IL-12 and IL-23 subunit *IL12B* in COVID-19, suggesting altered capacity for T cell polarisation (**Fig. 2E**). In peripheral blood, C1QA/B/C^+^CD16^+^ monocytes expressed the highest amount of Type 1 IFN response genes compared to all peripheral blood myeloid cells (**Fig. 2F**). We detected minimal TNF- or IL6-mediated JAK-STAT signaling pathway activation in circulating monocytes and DCs but this was upregulated by COVID-19 BAL mononuclear phagocytes (**Fig. 2F**).

Coagulation abnormalities and monocyte-platelet aggregates have been previously reported in COVID-19 patients^31,32^ and we observe an expansion of platelets associated with disease severity (**Fig. 1C**). This led us to investigate the receptor-ligand interactions predicted to mediate monocyte-platelet interactions using CellPhoneDB, which identified ICAM1 interactions on platelets with CD11a-c/CD18 primarily on C1QA/B/C^+^CD16^+^ monocytes and CD16^+^ monocytes (**Fig. 2G**). This is accompanied by increased expression of surface proteins indicative of platelet activation (**Fig. 2H**).

Our large dataset (850,100 PBMCs) allowed us to interrogate rare populations, including the HSPC compartment. To this end, we selected all cells in clusters with significant expression of the HSPC marker CD34, which resulted in a total of 3,085 HSPCs, following removal of minor clusters co-expressing mature lineage markers. Leiden clustering and UMAP visualisation resulted in a cloud-like representation with closely attached clusters, consistent with a stem/progenitor landscape previously described for bone marrow HSPCs^33^ (**Fig. 2I, Extended Data 3C**). Absence of CD38 mRNA and protein expression marks the most immature cells within the CD34 compartment, while expression of markers such as *GATA1, MPO* and *PF4* characterises distinct erythroid, myeloid and megakaryocytic progenitor populations (**Fig. 2I**). Accordingly, we were able to annotate six transcriptional clusters as CD34^+^CD38^-^ HSPCs, CD34^+^CD38^+^ early progenitor HSPCs, and CD34^+^ CD38^+^ erythroid, megakaryocytic and myeloid progenitors as well as a small population distinguished by the expression of genes associated with cell cycle (S-phase) (**Fig. 2I**).

Following stratification by disease severity, the most noteworthy observation was that the megakaryocyte progenitors were essentially absent in healthy and asymptomatic individuals, but comprised approximately 5% of CD34^+^ cells in mild, moderate, severe and critical patients (**Fig. 2J**). Unlike the bone marrow, which contains rapidly cycling progenitors, the peripheral blood is not thought to constitute a site for haematopoiesis^34^, consistent with the low number of CD34^+^ cells expressing a cell cycle signature, which was furthermore restricted to genes associated with S-phase (**Fig. 2I**). Disease-associated alterations of the circulating CD34^+^ progenitor cells are therefore a likely reflection of COVID-19 mediated perturbation of the normal homeostatic functioning of the bone marrow haematopoietic stem/progenitor compartment.

In light of our earlier observations of platelet activation and enhanced C1QA/B/C^+^CD16^+^ monocyte-platelet interactions (**Figs. 2G-H**), the appearance of CD34^+^ megakaryocyte progenitors was of particular interest, as it suggested a rebalancing of the stem/progenitor compartment. The overall number of these megakaryocyte progenitors however was low, prompting us to seek additional evidence for reprogramming of immature haematopoiesis. To this end, we carried out differential gene expression analysis between the megakaryocyte, myeloid and erythroid progenitor clusters, and used the resulting gene lists to build gene signatures in order to interrogate early activation or priming of lineage-specific transcriptional programs in the most immature haematopoietic progenitor cell clusters (**Extended Data 3D**). This analysis showed activation of the megakaryocyte progenitor signature in both the CD38^-^ and CD38^+^ HSPC populations (**Fig. 2K**), with less pronounced effects seen with the erythroid and myeloid signatures (**Extended Data 3E**). Of note, the megakaryocyte signature was also strongly induced in the asymptomatic patients, which do not contain substantial numbers of CD34^+^ megakaryocyte progenitors in their peripheral blood. Our earlier observation of increased platelet activation within the context of normal platelet counts (**Fig. 2H, Extended Data 2B**) is therefore consistent with a model whereby exaggerated megakaryopoiesis may be compensating for peripheral platelet consumption in COVID-19 patients. Of note, our HSPC compartment analysis suggests that immature haematopoiesis is also affected in asymptomatic patients, but possibly through distinct differentiation and/or cell mobilisation processes. Taken together, our data suggest that alterations in the cellular composition and transcription programs of the stem/progenitor compartment contribute to the patho-physiological response to SARS-CoV-2 infection.

### T-lymphocytes and TCR changes

To further characterise T-lymphocytes during the infection, we re-clustered the T cell compartment and identified 15 clusters of CD4 T cells, CD8 T cells, and innate-like T cells including γδ T cells, NKT cells, and MAIT cells across sample collection sites, donors and disease severity groups (**Fig. 3A, Extended Data 4**). Our cell annotation is based on both RNA and protein expression of marker genes, as well as effector cytokines (**Figs. 3B-C**). In the CD4 T cell compartment, we identified naïve CD4 T cells, central memory T cells (CD4 CM), effector memory T cells (CD4 EM), activated CD4 T cells expressing IL-22 (CD4 IL22), Th1 cells, Th2 cells, Th17 cells, Treg cells, and circulating T follicular helper cells (cTfh). In the CD8 compartment, we found naïve CD8 T cells, effector/cytotoxic T cells (CD8 TE), and effector memory T cells (CD8 EM) (**Fig. 3A**).

**Figure 3:**
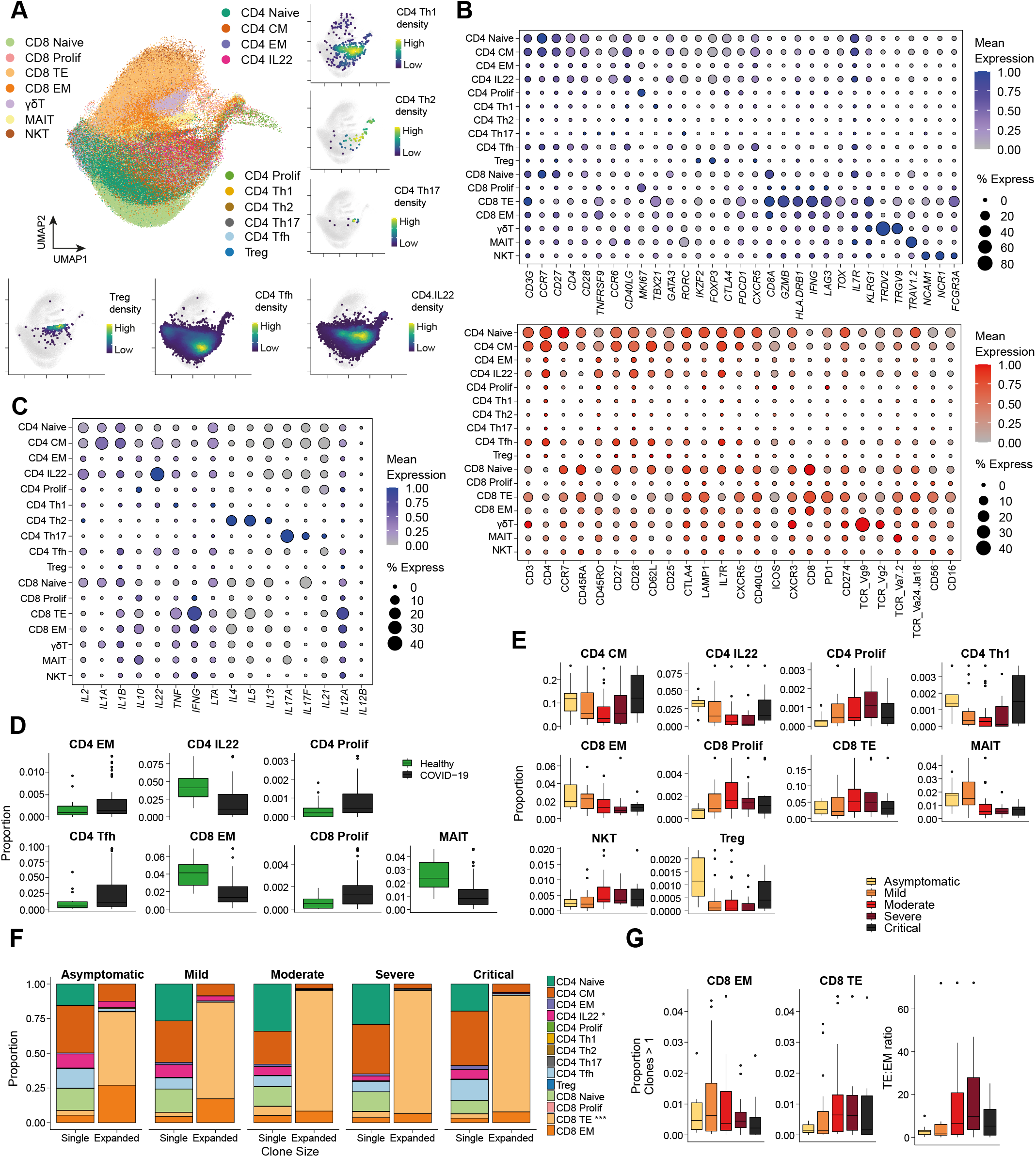
T lymphocytes. **A.** UMAP visualisation of T cells. Semi-supervised annotation of Louvain clusters based on gene expression shown and coloured by cell type. CM, central memory; EM, effector memory; TE, terminal effector; Th, T helper; Tfh, T follicular helper. Inset panels show the 2-dimensional kernel density estimates of select T cell types in UMAP space. **B**. Dot plots of gene expression (top; blue) and surface protein (bottom; red) expression for populations shown in **A**. where the colour is scaled by mean expression and the dot size is proportional to the percent of the population expressing the gene/protein, respectively. **C**. Dot plots of gene expression of cytokine genes for populations shown in **A**. where the colour is scaled by mean expression and the dot size is proportional to the percent of the population expressing the gene/protein, respectively. **D**. Box plots of cell type proportions that are differentially abundant between healthy donors and COVID-19 cases. Boxes denote interquartile range (IQR) with the median shown as horizontal bars. Whiskers extend to 1.5x the IQR; outliers are shown as individual points. **E**. Box plots of the proportion of cell types shown in **A**. separated by severity status. Only cell types showing trends of changes with respect to severity status are shown here. Boxes denote interquartile range (IQR) with the median shown as horizontal bars. Whiskers extend to 1.5x the IQR; outliers are shown as individual points. **F**. Bar plots showing the frequency of clonal T cells by severity. Expanded clones denote TCR clonotypes observed more than once. Stars in key indicate significance after multiple testing correction (Logistic regression; \**P* < 0.05, \*\**P* < 0.01, \*\*\**P* < 0.001). **G**. Box plots of the proportion of clonally expanded effector memory CD8 T cells (left), effector CD8 T cells (middle), and the ratio of effector CD8 T cells to effector memory CD8 T cells (right). Boxes denote interquartile range (IQR) with the median shown as horizontal bars. Whiskers extend to 1.5x the IQR; outliers are shown as individual points.

Cellular composition of the T cell compartment varied between the healthy and infected groups (**Fig. 3D)**. Notably, based on their relative proportions and differential abundance testing (FDR 10%), we found activated CD4 expressing IL-22, circulating Tfh cells, Th1 cells, Treg cells, CD8 EM cells, and MAIT cells relatively enriched in patients with asymptomatic and mild infection phenotype, with NKT, proliferating CD8 and CD4, and CD8 TE cells enriched in patients with more severe phenotypes (**Fig. 3E, Extended Data 5A-B**). Moreover, we observed multiple cell populations that displayed non-linear differences across severity phenotypes (proliferating CD4 & CD8, CD8 TE, CD4 Th1, CD4 Th17, CD4 CM, IL-22^+^ CD4, Treg), illustrating the complex compositional changes to peripheral T cells that occur with COVID-19 severity (**Fig. 3E, Extended Data 5B**). Interestingly, the enrichment of Treg cells and IL-22 expressing CD4 T cells in the patients with less severe disease (asymptomatic & mild) could be associated with immuno-regulatory and tissue-protective responses that may restrict immunopathology (**Fig. 3E**) as IL-22 was previously shown to be involved in tissue protection in influenza A virus infection^35^, and associated with low viral load in the lung parenchyma of COVID-19 patients^36^. The enrichment of proliferating CD4^+^ and CD8^+^ T cells, which also express some exhaustion marker genes (*LAG3, TOX*), could account for the previous observation of increased expression of exhaustion markers on CD8^+^ T cells in patients with severe disease^17^.

To investigate T cell phenotype beyond differential abundance of T cell subsets, we performed differential gene expression analysis across disease severity (FDR 1%) followed by gene set enrichment analysis (GSEA) in each cell type and found enrichment of pathways associated with inflammation and T cell activation across multiple T cell subsets, including *IL-2/STAT5* signaling, *mTORC1* signaling, inflammatory response, interferon gamma response, and *IL-6/JAK/STAT3* signaling (**Extended Data 5C**). Increases in activation and cytotoxic phenotype in the T cells from COVID-19 patients stimulated *ex vivo* with SARS-CoV-2 peptide was confirmed independently by protein expression of CD137 and CD107α using flow cytometry (**Extended Data 5D**).

Next, we interrogated TCR clonality and the relative proportions of specific T cell subsets within clonally expanded T cells in different disease groups (**Fig. 3F)**. As expected, among the COVID-19 patients, effector CD8 T cells were the most clonally expanded, with enriched large clone sizes, across different disease groups, and their relative proportion increased with disease severity (**Figs. 3F-G, Extended Data 5E-F**). Conversely, the relative proportion of clonally expanded effector memory CD8 T cells decreased in patients with more severe disease (**Figs. 3F-G**). The ratio of effector CD8 T cells to effector memory CD8 T cells correlated with disease severity (**Fig. 3G**), suggesting that CD8 T cell differentiation outcome may contribute to both anti-viral protection and immunopathology. This could be a result of the degree of inflammation set by innate immunity in the first instance, resulting in biased CD8 differentiation into antigen-specific short-lived effector CD8 T cells (equivalent to CD8 effector T cells in this study) *versus* memory precursor effector CD8 T cells (equivalent to CD8 effector memory T cells in this study), as previously reported in animal models^37^.

### B-lymphocytes and BCR changes

Re-clustering of B and plasma cells in isolation identified 9 clusters that were annotated according to canonical marker expression (**Figs. 4A-B**), and appropriate enrichment of previously published transcriptional signatures (**Extended Data 6A**). This included immature, naïve, switched and non-switched memory B cells, and a cluster of cells that enriched for markers previously described in exhausted memory B cells^38,39^ (**Figs. 4A-B, Extended Data 6A**). We also found a large population of CD19/CD20-negative plasmablasts, with high expression of the proliferation marker *MKI67*, as well as IgM^+^, IgG^+^, and IgA^+^ plasma cells (**Figs. 4A-B**). In patients with symptomatic COVID-19, there was a significant expansion of plasmablasts and plasma cells compared with healthy controls and LPS-treated subjects (**Fig. 4C, Extended Data 6B**). Notably, this phenomenon was less evident in COVID-19 patients with asymptomatic disease. IgG plasma cells, in particular, were expanded in symptomatic COVID-19 compared with other groups, and the magnitude of this expansion increased with worsening disease severity from mild to severe disease but surprisingly, was less evident in patients with critical disease (**Fig. 4C, Extended Data 6B**). When considering plasmablasts and plasma cells using the V(D)J data, IgA^+^ cells were expanded in patients with asymptomatic COVID-19 (**Fig. 4D**), suggestive of an effective mucosal humoral response in this patient group. This is paralleled by the greatest expansion of circulating Tfh cells in asymptomatic patients and underlined by the strong correlation between cTfh cells with plasma cells in asymptomatic/mild patients (**Fig. 3E, 4E, Extended Data 5A-B**), suggesting a potential contribution of coordinated T cells/B cells response to effective humoral anti-viral protection in these patients that is lost in severe and critical disease. This is consistent with previous findings relating to the requirement of Tfh cells for optimal antibody responses and high-quality neutralising antibodies in viral infection^40^.

**Figure 4:**
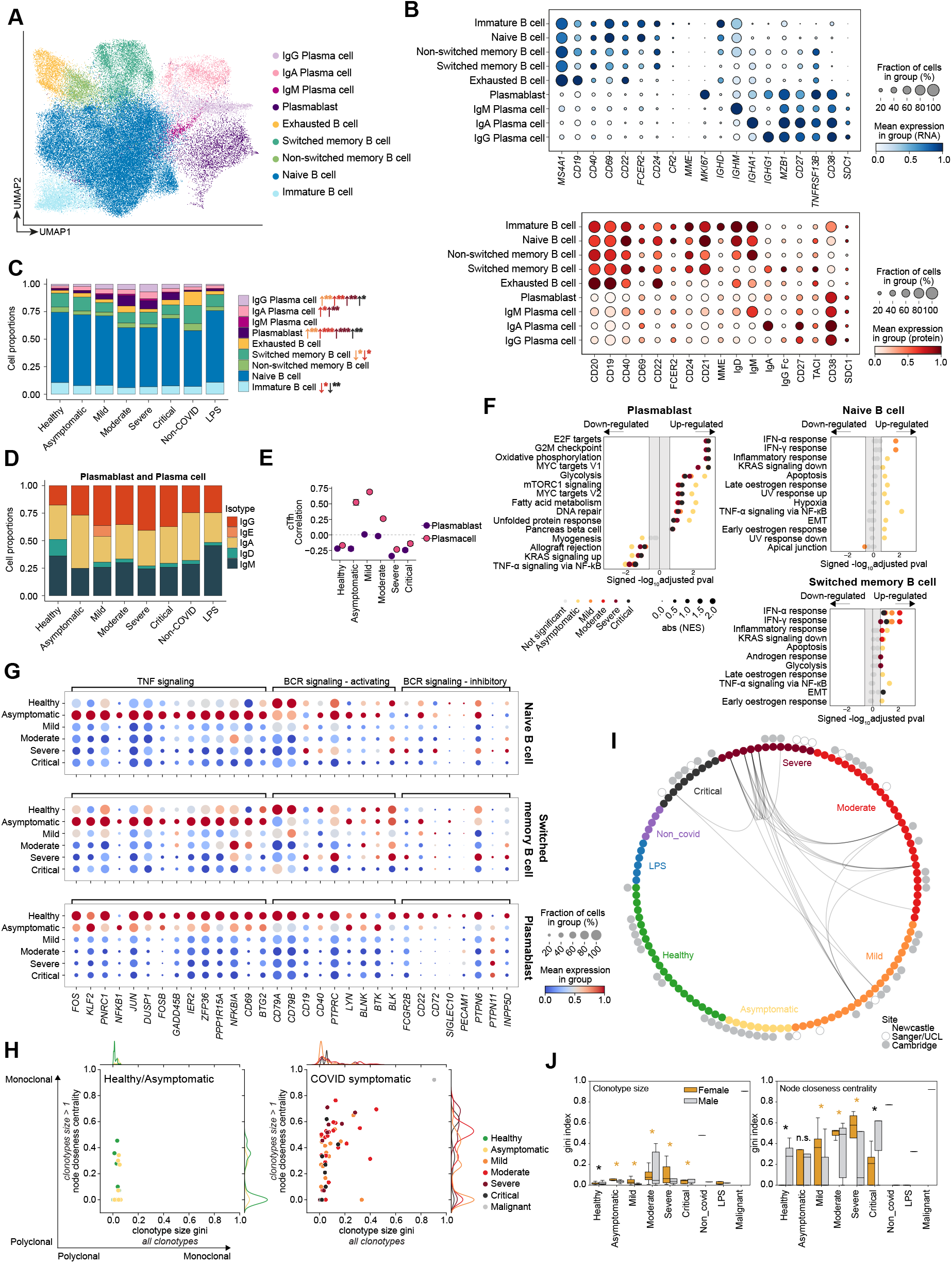
B lymphocytes. **A.** UMAP visualisation of 74,019 cells in the B cell lineage and coloured by cell type identified from clustering on the gene expression data. **B**. Dot plots of gene expression (top; blue) and surface protein (bottom; red) expression for populations shown in **A**. where the colour is scaled by mean expression and the dot size is proportional to the percent of the population expressing the gene/protein, respectively. **C**. Bar plot of the mean proportion of cell types shown in **A**. separated by severity status. Stars in key indicate significance (Kruskal-Wallis; \**P* < 0.05, \*\**P* < 0.01, \*\*\**P* < 0.001), arrows represent if proportional change is up or down and colour represents COVID-19 severity state. **D**. Bar plot showing the mean proportion of plasmablast and plasma cells expressing IgA, IgD, IgE, IgG or IgM, based on V(D)J information, separated by severity status. **E**. Co-ordinated changes between Tfh and B cells assessed by differential correlation analysis (empirical *P* ≤ 0.05). Shown is the Pearson correlation (+/-bootstrap standard error) between Tfh proportions and plasmablast or plasma cell (combined) according to disease severity (only significant trends are shown). **F**. GSEA of pathways from MSigDB hallmark signatures in naive B cells, switched memory B cells and plasmablast for asymptomatic/symptomatic COVID versus healthy. Size of circles indicate (absolute) normalised enrichment score (NES) and colours indicate the severity status. Pathways were considered statistically significant if *P* < 0.05 and FDR < 0.25 (denoted by coloured dots outside the middle grey zone). EMT, Epithelial-mesenchymal transition. **G**. Dot plots representing the expression of genes coding for TNF signalling molecules, activating and inhibitory BCR signaling molecules in naive B cells, switched memory B cells and plasmablast separated by severity status in the rows. Size of circles indicate percent of cells expressing the gene and increasing colour gradient from blue to white to red corresponds to increasing mean expression value (scaled from zero to one across status per gene). **H**. Scatter plot of clonotype size by node closeness centrality gini indices. Each dot represents the gini indices of an individual coloured by severity status. Gini indices were computed for all clonotypes on the x-axis and for clonotypes with > 1 cell on the y-axis (see methods for details). Marginal histograms indicate the distribution of samples in a given severity status along the axes. **I**. BCR overlap incidence plot. Nodes in the inner ring represent individual donors/patients, coloured by severity status, and edges indicate if at least 1 clonotype is shared between two individuals (at least 1 cell in each individual displays an identical combination of heavy and light chain V- and J-gene usage with allowance for somatic hypermutation at the CDR3 junctional region). Nodes in the outer ring indicate the site from which samples were collected (solid grey: Cambridge; grey outline: Sanger; unmarked: Newcastle) **J**. Clonotype size (left panel) and node closeness centrality gini indices (right panel) separated by gender. Statistical tests were performed with non-parametric Mann-Whitney U test between the gender groups within each severity status and were considered statistically significant if Benjamini-Hochberg corrected *P* < 0.05 (denoted by *; n.s. denotes not significant). Colour of asterisks indicates which gender group displays a higher mean gini index (yellow: female; grey: male).

To interrogate the effect of COVID-19 infection on humoral immune cells beyond differential expansion of subsets, we performed GSEA in each cell type. *Interferon alpha response* and *interferon gamma response* pathway genes were enriched in all B cell subsets in COVID-19 patients, but this response was generally more marked in patients with asymptomatic or mild disease, and attenuated in severe and critical disease (**Fig. 4F, Extended Data 6C**). The magnitude of type 1 interferon transcriptional response in B cells mirrored serum IFN-α levels, which were highest in patients with mild disease (**Extended Data 2H**), suggesting that the low expression of IFN response genes in B cells in severe or critical disease does not reflect an inability of B cells to respond to IFN-α, but rather attenuation of IFN-α. This may be because the initial anti-viral response has waned in patients with severe or critical disease or because these patients fail to sustain adequate IFN-α production by myeloid cells and pDCs following symptom onset as previously reported^13^. Longitudinal sampling would be required to distinguish these two possibilities.

In asymptomatic patients, *TNFA signalling via NF-kB* pathway genes were also enriched in immature, naïve and switched memory B cells, but decreased in immature B cells and plasma cells in critical and severe disease (**Fig. 4F, Extended Data 6C**). Assessment of the leading-edge genes in this pathway demonstrated their markedly higher expression in all B cell and plasmablast/cell subsets in asymptomatic COVID-19 patients compared with those with symptomatic disease (**Fig. 4G, Extended Data 6D**). TNFa was barely detectable in COVID-19 serum samples and highest in patients with moderate disease (**Extended Data 1E**), suggesting that another cytokine e.g. IL-6 or stimulus may be responsible for NF-kB activation in asymptomatic COVID-19 patients.

Hypoxia pathway genes were enriched in immature and naïve B cells only in asymptomatic patients (**Fig. 4F, Extended Data 6C**). Since these individuals are unlikely to be hypoxic (given their lack of symptoms) we postulated that this signature may reflect another hypoxia-inducible factor (HIF) activating stimulus, which includes B cell receptor (BCR) cross-linking^41^. We assessed the expression of genes associated with BCR activation, such as *CD79A/B*, and downstream kinases such as *BTK* in B cell subsets. Overall, BCR activation-associated genes were most highly expressed in B cells in healthy control cells, followed by asymptomatic COVID-19 patients, with lower expression observed in all symptomatic COVID-19 groups (**Fig. 4G, Extended Data 6D**). BCR activation threshold is also modulated by immune tyrosine inhibitory motif (ITIM)-containing receptors that recruit phosphatases, increasing the activation threshold of B cells^42^. BCR inhibitory gene expression was limited, but *CD22* was detectable across B cell subsets in asymptomatic COVID-19, whilst *FCGR2B, CD72* and *PTPN6* expression was evident in severe COVID-19 B cells (**Fig. 4G, Extended Data 6D**). Together, this analysis suggests that B cells in asymptomatic COVID-19 patients and those with mild disease have a more pronounced response to interferons, increased NF-kB activation, and a higher expression of genes associated with BCR activation signaling, suggesting a potential for greater BCR activation. This may indicate that more avid responses early in disease prevent progression to a more severe phenotype, or may merely reflect the immune response in the early phase of the disease. Longitudinal analysis of patient samples would be required to address this question.

Following activation, B cells differentiate into antibody-producing plasma cells, accompanied by a progressive increase in oxidative metabolism^43,44^. We observed differences in metabolic gene pathway expression in plasmablasts and plasma cells between disease severity categories, with enrichment of *oxidative phosphorylation* pathway genes in critical and severe disease, but increase in *glycolysis* pathway genes in asymptomatic patient plasmablasts (**Fig. 4F, Extended Data 6D**).

We next assessed BCR clonality using *dandelion*, a novel single cell BCR-sequencing analysis package (see methods), and found significantly more clonal expansion in symptomatic COVID-19 patients compared with those with asymptomatic disease or healthy controls (**Fig. 4H, Extended Data 7A**). Expanded clonotypes were found across all major cell types with larger clonotypes primarily present in plasmablast/plasma cell clusters (**Extended Data 8A-B**). Within the expanded clonotypes, there was some evidence of class switching within symptomatic COVID groups but not in the asymptomatic/healthy (**Extended Data 8C**). Some related BCRs were present in different individuals, with more incidence of V-, J-gene usage and related amino acid sequences of heavy and light chain CDR3s observed in patients with severe or critical disease, and in patients within one of the clinical centres (Newcastle) (**Fig. 4I**), which could arise due to local variants of the virus driving expansion of specific B cell clones. We note that none of these related BCRs were found to be expanded in the individuals which was expected as only a relatively small number of B cells per individual were sampled. It would have been extremely unlikely to find exactly matching heavy- and light-chain sequences across different individuals (even when allowing for somatic hypermutation variation) given the expected low coverage that arises from a small number of cells (relative to bulk BCR sequencing). Finally, we observed disproportionate distribution in clonotype size, whether considering expanded or all clonotypes, and increased BCR mutation between male and female patients, with greater levels of both in females compared with males (**Fig. 4J, Extended Data 7B**). These differences in clonal expansion of B cells are consistent with previous reports of worse outcomes in COVID-19 in males^45,46^.

We summarise the immunological cellular and molecular profiles observed in our study highlighting known and new discoveries as well as the distinguishing features of asymptomatic and milder disease from severe and critical disease (**Fig. 5**). Future longitudinal studies may enable us to distinguish if the distinct responses in asymptomatic and milder disease prevent progression to severe phenotypes.

**Figure 5:**
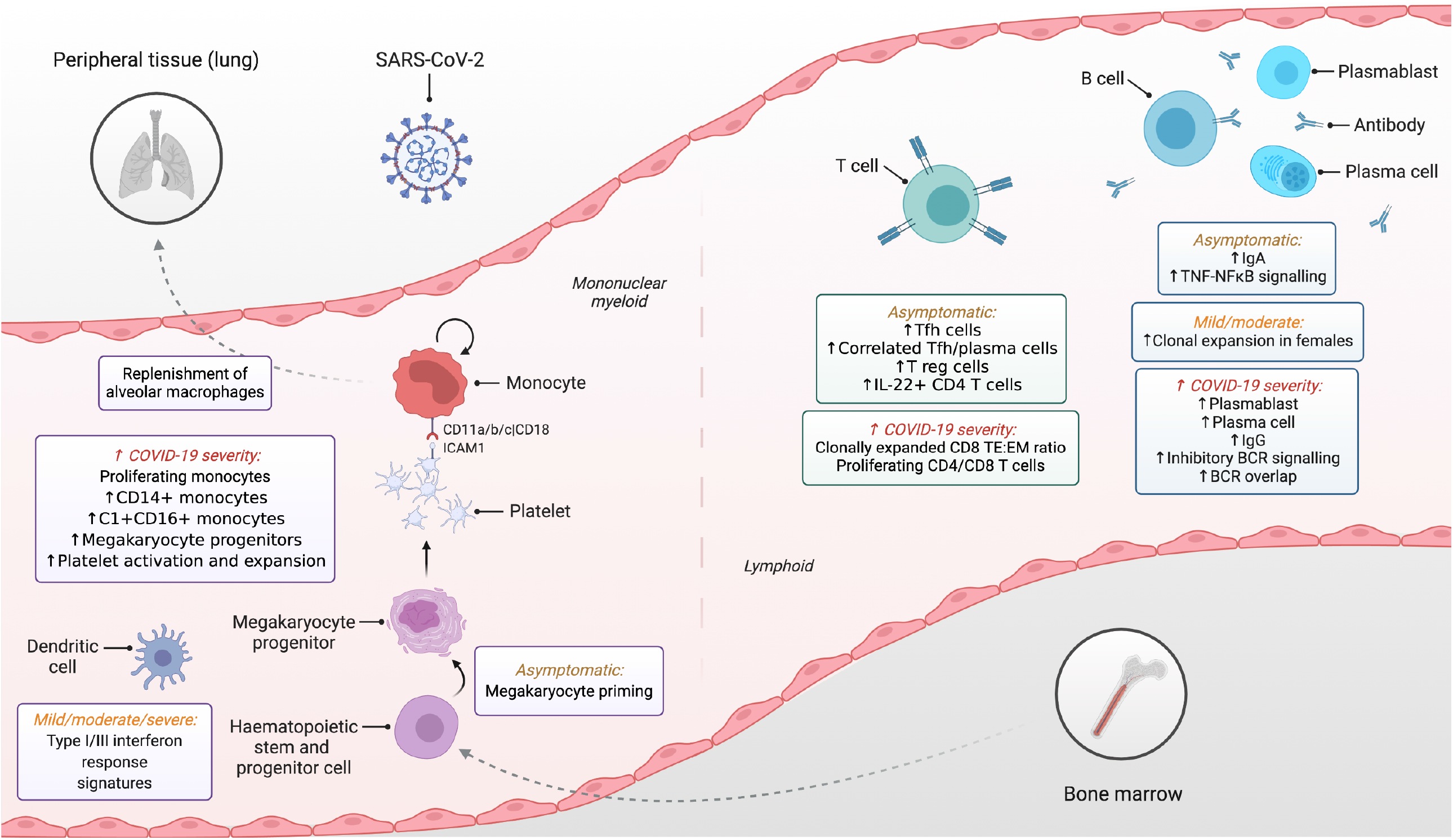
Integrated framework of mononuclear cell immune response in blood. Schematic illustration of study highlights. Created with Biorender.com. BCR, B cell receptor; EM, effector memory; TE, terminal effector; Tfh, T follicular helper; Th, T helper; T reg, regulatory T cell

## Discussion

Our cross-sectional multi-omics peripheral blood mononuclear cell survey of ∼130 COVID-19 patients and controls across three UK centres revealed several new insights into COVID-19 pathogenesis. Firstly, peripheral blood monocytes and DCs exhibit an interferon response to infection and replenish peripheral tissue mononuclear phagocytes such as alveolar macrophages. Secondly, the initial peripheral tissue inflammation and systemic response to COVID-19 is accompanied by altered haematopoiesis that is mirrored in the peripheral circulation evidenced by megakaryocyte-primed gene expression in the earliest CD34^+^CD38^-^ HSPCs, exaggerated megakaryopoiesis and platelet activation. CD1QA/B/C^+^CD16^+^ monocytes co-express receptor:ligands predicted to interact with platelets, supporting their intertwined role in tissue thrombosis reported in COVID-19.

We reveal a balance in protective versus immunopathogenic adaptive immune responses in COVID-19 patients. In patients with less severe disease, we found enrichment of circulating Tfh cells, which were previously shown to also be involved in SARS-CoV-2 infection^47,48^ and Th1 cells, which could also confer anti-viral protection^49,50^. Our findings suggest that an imbalance in CD8 T cell differentiation, including the overexpansion of CD8 effector T cells which likely include antigen-specific short-lived effector cells, could lead to uncontrolled inflammation and immunopathology. Whether the reduced proportion of clonally expanded CD8 effector memory T cells could lead to impaired memory responses in patients with more severe disease remains an open question to be further investigated.

Similarly, in B cells, expansion of plasmablasts and plasma cells is less evident in critical than in moderate and severe patients. This response is paralleled by the Tfh profile in COVID-19 patients and is consistent with post-mortem observations showing a lack of GCs in lymph nodes and spleen in patients with fatal COVID-19 and a decrease in Tfh^48^. We observe a diminished IFN-α response in critical and severe patients’ B cell compartments, further emphasising a critical role of these responses in outcomes, as previously reported in COVID-19 patients with anti-type I IFN antibodies^51^. The presence of common BCRs in samples from one geographical region could reflect local differences in viral strain, with increasing awareness of how viral mutations may influence outcomes, as has been shown to be clinically important for viral transmission with the B.1.1.7 strain^52,53^.

Our cross-sectional study demonstrates valuable new insights from multi-omics profiling of peripheral blood as a window to understand peripheral tissue inflammation, as well as bone marrow and systemic responses to acute COVID-19 infection. Our large datasets and web portal provide a foundational resource on COVID-19 for the research and clinical communities.

## Data Availability

The dataset from our study can be explored interactively through a web portal: https://covid19cellatlas.org. The data object, as a h5ad file, can also be downloaded from the portal page. The processed data is available to download from Array Express using accession number E-MTAB-10026.

## Acknowledgements

We acknowledge assistance from Peter Vegh, James Fletcher and David Dixon to the IV-LPS study and funding from the Wellcome Human Cell Atlas Strategic Science Support (WT211276/Z/18/Z). We acknowledge Bertrand Yeung and Kristopher “Kit” Nazor from Biolegend for helpful discussions in the optimisation of the CITE-Seq protocol. We acknowledge Rachel Queen and Rafiqul Hussain from Newcastle University Genomics Core Facility for technical assistance. We acknowledge Martin Prete for assistance with online data hosting and interactivity. We thank NIHR BioResource volunteers for their participation, and gratefully acknowledge NIHR BioResource centres, NHS Trusts and staff for their contribution. We thank the National Institute for Health Research, NHS Blood and Transplant, and Health Data Research UK as part of the Digital Innovation Hub Programme. The views expressed are those of the author(s) and not necessarily those of the NHS, the NIHR or the Department of Health and Social Care. M.H. is funded by Wellcome (WT107931/Z/15/Z), The Lister Institute for Preventive Medicine and Newcastle NIHR Biomedical Research Centre (BRC); B.G. is funded by Wellcome (206328/Z/17/Z), MRC (MR/S036113/1), and the Aging Biology Foundation. S.A.T. is funded by Wellcome (WT206194), ERC Consolidator and EU MRG-Grammar awards. Z.K.T. and M.R.C. are supported by a Medical Research Council Human Cell Atlas Research Grant (MR/S035842/1). J.C.M. is supported by core funding from Cancer Research UK (C9545/A29580) and from the European Molecular Biology Laboratory (EMBL). M.R.C. is supported by a Versus Arthritis Cure Challenge Research Grant (21777), and an NIHR Research Professorship (RP-2017-08-ST2-002). H.W.K. is supported by Sir Henry Wellcome PostDoctoral fellowship Wellcome Trust (213555/Z/18/Z). K.B.M acknowledges funding from the Chan Zuckerberg Initiative (grant 2017-174169) and from Wellcome (WT211276/Z/18/Z and Sanger core grant WT206194). M.Z.N. acknowledges funding from a UKRI Innovation/Rutherford Fund Fellowship allocated by the MRC and the UK Regenerative Medicine Platform (MR/5005579/1 to M.Z.N.). M.Z.N. and K.B.M. have been funded by the Rosetrees Trust (M944). K.B.W. is funded by University College London, Birkbeck MRC Doctoral Training Programme. J.L.B. acknowledges funding from the MRC and the UK Regenerative Medicine Platform (MR/5005579/1). M.Y. is funded by The Jikei University School of Medicine. K.F.B is funded by an NIHR Clinical Lectureship (CL-2017-01-004). N.M. is supported by a DFG Research Fellowship (ME 5209/1-1). C.J.A.D is funded by Wellcome (211153/Z/18/Z) and acknowledges support from the Barbour Foundation. This work was partly funded by UKRI/NIHR through the UK Coronavirus Immunology Consortium (UK-CIC).

## Author Contributions

M.H., B.G., S.T., J.M., M.R.C., M.Z.N., K.M. conceived the study. A.J.S., A.J.R. conceived the IV-LPS study. C.J.A.D., K.S., P.L., M.Z.N., E.K., A.dW., A.Sai., A.Sal., S.M.J., A.T.H., K.F.B., I.C.D.SvdL., L.C.S.G., A.S.B., A.S.G., L.B. recruited patients, collected samples and clinical metadata. E.S., R.A.B., F.G., J.S., R.P.P., K.B.W., M.Y., J.L.B., N.M., F.J.C. isolated PBMCs. R.A.B., E.S., K.B.W., M.Y., J.L.B., N.M., F.J.C-N. performed 10x and CITE-seq. E.S., J.E., K.K. prepared sequencing libraries. E.P., J.C., P.C. conducted the sequencing. G.R., Z.K.T., K.B., M.M., W.S., N.K., S.vD., V.K., N.H., R.L., K.P., E.D. analysed the data. M.H., B.G., S.T., J.M., M.R.C., M.Z.N., K.M., G.R., Z.K.T., K.B., M.M., W.S., N.K., R.A.B., E.S., L.J., S.W., J.S.S. interpreted the data. J.S.S. performed flow cytometry. E.S. performed multiplex cytokine analysis. M.H., B.G., S.T., M.R.C., J.M., E.S., G.R., R.B., L.J., B.O., M.M., K.B., N.K., W.S., M.Z.N. wrote the manuscript. N.M., LC.S.G., S.W., K.F.B., F.M., C.W., J.M.C., H.W.K., S.H., E.L., K.B.M., K.S., edited the manuscript. Z.K.T., H.W.K. developed software (*dandelion*). J.McG., D.H. developed the web portal.

## Code availability

All data analysis scripts are available on https://github.com/scCOVID-19/COVIDPBMC

**Extended Data 1**

**A**. Scatter plot displaying the total number of gene counts per sample from each site. **B**. UMAPs from **Fig. 1B** coloured by site. **C**. Boxplot of kBET results calculated both before and after batch correction with Harmony for each cluster in **Fig. 1B** kBET statistic calculating using patient ID as the batch factor. **D**. Dot plots of 5’ gene expression (top; blue) and surface protein (bottom; red) expression for populations shown in **Fig. 1A** where the colour is scaled by mean expression and the dot size is proportional to the percent of the population expressing the gene/protein, respectively. **E**. Tile plot showing percentage concordance between COVID-19 PBMC annotation (y-axis) and Azimuth annotation (x-axis) (https://satijalab.org/azimuth/).

**Extended Data 2**

**A**. Volcano plots showing results of differential abundance testing. Hypothesis testing was performed using quasi-likelihood F-test comparing healthy controls to cases for linear trends across disease severity groups (healthy > asymptomatic > mild > moderate > severe > critical). Differentially abundant cell types were determined using a 10% false discovery rate (FDR) and marked (*). Hypothesis testing was performed using quasi-likelihood F-test comparing healthy controls to cases. Differentially abundant cell types were determined using a 10% false discovery rate (FDR). **B**. Box and whisker plots showing blood counts for Newcastle data grouped by severity status. Dotted lines and green area mark the normal ranges for each. Kruskal-Wallis with Dunn’s post hoc; \**P* < 0.05, \*\**P* < 0.01. **C**. Forest plot showing the standard deviation of each clinical/technical factor estimated by the Poisson generalised linear mixed model. The error bars show the standard error estimated from the Fisher information matrix (see **Supplementary Note 1** for more details). SD, standard deviation. **D**. Box plots displaying the duration of COVID-19 symptoms from the onset grouped by severity status. **E**. Volcano plots showing results of differential abundance testing according to time since symptom onset. Differentially abundant (FDR 10%) points are shown in red and labelled by cell type as in **Figure 1A. F**. Correlated log fold-changes of cell type abundance changes as a function of symptom duration with (x-axis) and without critically ill patients (y-axis). **G**. Heat map displaying fold change over healthy (left) and dot plot of gene expression where the colour is scaled by mean expression and the dot size is proportional to the percent of the population expressing the gene (right) for genes associated with COVID-19 identified in a recent GWAS study^25,26^ for the cell populations in **Fig. 1B. H**. Heat map displaying normalised values of cytokine, chemokine and growth factors in serum of patients with COVID-19.

**Extended Data 3**

**A**. Dot plots of gene expression of C1 complement components for cells in **Fig. 1B** where the colour is scaled by mean expression and the dot size is proportional to the percent of the population expressing the gene. **B**. Dot plots of gene expression of a recently published BAL dataset (accession number GSE145926^29^) for genes in **Fig. 2A** where the colour is scaled by mean expression and the dot size is proportional to the percent of the population expressing the gene. **C**. Heatmap of differentially expressed genes between megakaryocyte, myeloid and erythroid progenitor clusters. MK, megakaryocyte; My, myeloid. **D**. Bar charts displaying enrichment of an erythroid signature (top) and a myeloid signature (bottom) found in CD34^-^CD38^-^ (left) and CD34^+^CD38^+^ HSPCs (right), separated by severity.

**Extended Data 4**

**A**. UMAP visualisation of T cells separated by sources of donors. **B**. UMAP visualisation showing 2-dimensional kernel density estimates of each T cell type in UMAP space. **C**.-**E**. UMAP visualisation of T cells coloured by gender (**C**.), disease severity status (**D**.) and age (**E**.).

**Extended Data 5**

Box plots showing the proportion of cell types shown in **Fig. 3A** separated by severity status. Volcano plots showing results of differential abundance testing. Cell type abundance counts were modelled either comparing healthy vs. COVID-19 case, or as a function of disease severity. Hypothesis testing was performed using quasi-likelihood F-test comparing healthy controls to cases, or for either a linear or quadratic trend across disease severity groups (asymptomatic > mild > moderate > severe > critical). Differentially abundant cell types were determined using a 10% false discovery rate (FDR). **C**. Gene set enrichment (MSigDB Hallmark 2020) in each T cell type based on differential gene expression (DGE) analysis was performed across COVID-19 disease severity groups, ordered from healthy > asymptomatic > mild > moderate > severe > critical. Statistically significant DE genes were defined with FDR < 0.01. Significant enrichments were defined with 10% FDR. **D**. Bar plots showing percent (mean +/-SEM) of CD3^+^CD4^+^ (blue) and CD3^+^CD8^+^ (green) T cells expressing CD107a (left) and CD137 (right) in response to SARS-CoV-2 S peptide stimulation. Significance determined using Kruskal-Wallis with Dunn’s post-hoc; \**P* < 0.05, \*\**P* < 0.01. **E**. Box plots showing clone size distribution for each T cell subset separated by severity status. **F**. Box plots slowing clonal diversity for each T cell subset separated by severity status.

**Extended Data 6**

**A**. Heatmap of mean gene set enrichment scores of (top) adult peripheral blood B cell signatures^38^ and (bottom) Human cell atlas bone marrow B cell signatures^54^. Enrichment scores were calculated using *scanpy*’s *tl*.*score_genes* function, tabulated as the mean of each cell type. Row enrichment value is scaled from 0 to 1 and presented as an increasing gradient from purple, blue, green to yellow which corresponds to increasing mean enrichment score. **B**. (Top) Kruskal-Wallis test results with Benjamini-Hochberg false discovery correction for cell type proportion differences in plasmablast and plasma cells between severity statuses. Significance is denoted by \**P* < 0.05; \*\**P* < 0.01; \*\*\**P* < 0.001. (Bottom) Cell type abundance counts were modelled as a function of disease severity. Hypothesis testing was performed using quasi-likelihood F-test comparing asymptomatic to symptomatic covid, for either a linear or quadratic trend across disease severity groups (asymptomatic > mild > moderate > severe > critical). Differentially abundant cell types were determined using a 10% false discovery rate (FDR). **C**. GSEA of pathways from MSigDB v7.2 hallmark signatures in immature B cells, non-switched memory B cells, ‘exhausted’ B cells and plasma cells for asymptomatic/symptomatic COVID versus healthy. Size of circles indicate (absolute) normalised enrichment score (NES) and colours indicates the severity status. Pathways were considered statistically significant if *P* < 0.05 and FDR < 0.25 (denoted by coloured dots outside the middle grey zone). EMT, Epithelial-mesenchymal transition. **D**. Dot plots of TNF signalling molecules, activating and inhibitory BCR signaling molecules (5’ gene expression data) in immature B cells, non-switched memory B cells, ‘exhausted’ B cells and plasma cells separated by severity status in the rows. Size of circles indicate percent of cells expressing the gene and increasing colour gradient from blue to white to red corresponds to increasing mean expression value (scaled from zero to one across status per gene).

**Extended Data 7**

**A**. Single-cell BCR network plots for each severity status coloured by heavy chain isotype class (IgM, IgD, IgA, IgE, or IgG). Each circle/node corresponds to a single B cell with a corresponding set of BCR(s). Each clonotype is presented as a minimally connected graph with edge widths scaled to 1/*d*+1 for edge weight *d* where *d* corresponds to the total (Levenshtein) edit distance of BCRs between two cells. Size of nodes is scaled according to increasing node closeness centrality scores i.e. nodes that are highly central to a clonotype network will be larger. **B**. (Left) Scatter plot of clonotype/cluster size by vertex size gini indices computed from contracted BCR networks (identical nodes are merged and counted). Each dot represents the gini indices of an individual coloured by severity status. Gini indices were computed for all clonotypes on both x-y-axes (see methods for details). Marginal histograms indicate the distribution of samples in a given severity status along the axes. (Right, top) Cluster/clonotype size (contracted network) gini indices separated by gender. (Right, bottom) Vertex size (contracted network) gini indices separated by gender. Statistical tests were performed with non-parametric Mann-Whitney U test between the gender groups within each severity status and were considered statistically significant if Benjamini-Hochberg corrected *P* < 0.05 (denoted by *; n.s. denotes not significant). Colour of asterisks indicates which gender group display a higher mean gini index (yellow: female; grey: male).

**Extended Data 8**

**A**. UMAP visualisation of B cell lineage and coloured by clonotype size in the V(D)J data. Only expanded clonotypes are coloured (clonotype size > 2). **B**. Single-cell BCR network plots for each severity status coloured by assigned cell type. **C**. Single-cell BCR network plots for each severity status coloured by heavy chain isotype subclass (IgM, IgD, IgA1, IgA2, IgE, IgG1, IgG2, IgG3 or IgG4). Each circle/node corresponds to a single B cell with a corresponding set of BCR(s). Each clonotype is presented as a minimally connected graph with edge widths scaled to 1/*d*+1 for edge weight *d* where *d* corresponds to the total (Levenshtein) edit distance of BCRs between two cells. Size of nodes is scaled according to increasing node closeness centrality scores i.e. nodes that are highly central to a clonotype network will be larger.

**Supplementary Information Guide**

**Supplementary Table 1:** Patient metadata. Status summary is based on the WHO COVID-19 classification (WHO reference number: WHO/2019-nCoV/clinical/2020.5; http://www.who.int/publications/i/item/clinical-management-of-covid-19)” www.who.int/publications/i/item/clinical-management-of-covid-19). NA, not applicable. Not-known listed where information was unavailable. O2, supplemental oxygen via nasal cannulae, face mask or non-rebreathe mask. NIV, non-invasive ventilation under continuous (CPAP) or bi-level (BiPAP) positive airways pressure.

**Supplementary Table 2: CITE-seq panel**. List of Total-seq C antibodies, including clone and barcode.

**Supplementary Table 3:** Clinical whole blood counts for Newcastle samples. Number of cells x 10^9^/L of blood. WBC, white blood cells.

**Supplementary Table 4:** Concentration in pg/mL of 45 analytes measured in serum. <=0, below the limit of detection; * indicates anti-inflammatory cytokines.

**Supplementary Note 1:** Further information detailing the poisson linear mixed model for cell type composition analysis.

**Supplementary Note 2:** List of collaborators and their affiliations from the CITIID-NIHR COVID-19 BioResource.

## Methods

### Ethics and sample collection

#### Newcastle

Patients were consented under the Newcastle Biobank (REC 17/NE/0361, IRAS 233551) study and ethical governance. For the COVID-19 positive samples and healthy controls, peripheral blood was collected in EDTA tubes and serum separator tubes and processed within 4 h of collection.

For the IV-LPS control samples: Ethical approval was granted by a REC (17/YH/0021). Healthy volunteers gave informed, written consent. LPS was obtained from Clinical Center Reference Endotoxin (Lots 94332B1 donated by National Institute of Health, Bethesda, Maryland, USA) and injected intravenously as a bolus dose of 2 ng/kg. Blood samples were taken prior to IV LPS administration (baseline) and at 90 min, and 10 h post challenge. Venous blood was drawn from an 18g venous cannula and was collected into EDTA and serum separator tubes. Only samples from 90 min and 10 h were analysed in this study.

#### Cambridge

Study participants were recruited between 31/3/2020 and 20/7/2020 from patients attending Addenbrooke’s Hospital with a suspected or nucleic acid amplification test (NAAT) confirmed diagnosis of COVID-19 (including point of care testing (Collier et al., 2020; Mlcochova et al., 2020)), patients admitted to Royal Papworth Hospital NHS Foundation Trust or Cambridge and Peterborough Foundation Trust with a confirmed diagnosis of COVID-19, together with Health Care Workers identified through staff screening as PCR positive for SARS-CoV-2 (Rivett et al., 2020). Controls were recruited among hospital staff attending Addenbrooke’s serology screening programme, and selected to cover the whole age spectrum of COVID-19 positive study participants, across both genders. Only controls with negative serology results (45 out of 47) were subsequently included in the study. Recruitment of inpatients at Addenbrooke’s Hospital and Health Care Workers was undertaken by the NIHR Cambridge Clinical Research Facility outreach team and the NIHR BioResource research nurse team. Ethical approval was obtained from the East of England – Cambridge Central Research Ethics Committee (“NIHR BioResource” REC ref 17/EE/0025, and “Genetic variation AND Altered Leukocyte Function in health and disease - GANDALF” REC ref 08/H0308/176). All participants provided informed consent. Each participant provided 27 mL of peripheral venous blood collected into a 9 mL sodium citrate tube.

#### UCL/Sanger

Subjects 18 years and older were included from two large hospital sites in London, United Kingdom, namely University College London Hospitals NHS Foundation Trust and Royal Free London NHS Foundation Trust during the height of the pandemic in the United Kingdom (April to July 2020). Ethical approval was given through the Living Airway Biobank, administered through UCL Great Ormond Street Institute of Child Health (REC reference: 19/NW/0171, IRAS project ID 261511), as well as by the local R&D departments at both hospitals. At daily virtual COVID-19 co-ordination meetings suitable patients were chosen from a list of newly diagnosed and admitted patients within the preceding 24 h (based on a positive nasopharyngeal swab for SARS-CoV-2). Patients with typical clinical and radiological COVID-19 features but with a negative screening test for SARS-CoV-2 were excluded. Other excluding criteria included active haematological malignancy or cancer, known immunodeficiencies, sepsis from any cause and blood transfusion within 4 weeks. Maximal severity of COVID-19 was determined retrospectively by determining the presence of symptoms, the need of oxygen supplementation and the level of respiratory support. Peripheral blood sampling was performed prior to inclusion to any pharmacological interventional trials.

Samples were collected and transferred to a Category Level 3 facility at University College London and processed within 2 h of sample collection. Peripheral blood was centrifuged after adding Ficoll Paque Plus and PBMCs, serum and neutrophils separated, collected and frozen for later processing.

### Clinical status assignment

Clinical metadata was collected at the point of sample collection, including current oxygen requirements and location. This was used to assign disease severity status. Patients based on a ward and not requiring oxygen were defined as “Mild”. Patients outside of an intensive care unit (ICU) environment requiring oxygen were defined as “Moderate”. All patients on ICU and/or requiring non-invasive ventilation were defined as “Severe”. Patients requiring intubation and ventilation were defined as “Critical”. There were no patients in ICU that did not require supplemental oxygen.

### PBMC isolation and dead cell removal

#### Newcastle

PBMCs were isolated from blood samples using Lymphoprep (StemCell Technologies) density gradient centrifugation as per manufacturer’s instructions. Single cell suspensions were then washed with Dulbecco’s phosphate buffered saline (PBS) (Sigma) and frozen in 5-10 million cell aliquots in 90% (v/v) heat inactivated fetal calf serum (FCS) (Gibco) 10% (v/v) DMSO (Sigma Aldrich). On the day of the experiment the cells were thawed for 1 min, transferred to Wash buffer (PBS supplemented with 2% (v/v) FCS and 2 mM EDTA), and centrifuged at 500 g for 5 min. Resuspended cells were passed through a 30 μm filter and counted prior to live cell MACS enrichment with the Dead cell removal kit (Miltenyi Biotech) as per manufacturer’s instructions. Cell pellets were resuspended in microbeads and incubated at room temperature for 15 min. Each stained sample was passed through an LS column (Miltenyi Biotec) and rinsed with Binding buffer (Miltenyi Biotec) before centrifugation. Cell pellets were resuspended in Wash buffer and counted for CITE-seq antibody staining.

#### Cambridge

Peripheral blood mononuclear cells (PBMCs) were isolated using Leucosep tubes (Greiner Bio-One) with Histopaque 1077 (Sigma) by centrifugation at 800 g for 15 min at room temperature. PBMCs at the interface were collected, rinsed twice with autoMACS running buffer (Miltenyi Biotech) and cryopreserved in FBS with 10% DMSO. All samples were processed within 4 h of collection. Purified PBMCs were thawed at 37°C, transferred to a 50 mL tube and 10 volumes of pre-warmed thawing media (IMDM (Gibco 12440-053) with 50% (v/v) FCS (not heat inactivated; Panbiotech P40-37500) and 0.1 mg/mL DNaseI (Worthington LS002139)) were added slowly and dropwise, followed by centrifugation at 500 g for 5 min. The pellet was resuspended in 1 mL of FACS buffer (PBS (Sigma D8537) with 3% (v/v) heat-inactivated FCS) and viability of each sample was assessed by counting in an improved Neubauer chamber using Trypan blue. Pools of 4 samples were generated by combining 0.5 million live cells per individual (2 million live cells total). The pools were washed twice in FACS buffer (10 mL and 2 mL, respectively) followed by centrifugation for 5 min at 500 g. The pellet was then resuspended in 35 μL of FACS buffer and the viability of each pool was assessed.

#### UCL/Sanger

Peripheral whole blood was collected in EDTA tubes and processed fresh via Ficoll-Paque Plus separation (GE healthcare,17144002). The blood was first diluted with 5 mL 2 mM EDTA-PBS (Invitrogen, 1555785-038), before 10-20 mL of diluted blood was carefully layered onto 15 mL of Ficoll in a 50 mL falcon tube. If the sample volume was less than 5 mL, blood was diluted with an equal volume of EDTA-PBS and layered onto 3 mL Ficoll. The sample was centrifuged at 800 g for 20 min at room temperature. The plasma layer was carefully removed and the peripheral blood mononuclear cell (PBMC) layer collected using sterile Pasteur pipette. The PBMC layer was washed with 3 volumes of EDTA-PBS by centrifugation at 500 g for 10 min. The pellet was suspended in EDTA-PBS and centrifuged again at 300 g for 5 min. The PBMC pellet was collected and the cell number and viability assessed using Trypan blue. Cell freezing medium (90% FBS, 10% DMSO) was added dropwise to PBMCs slowly on ice and the mixture cryopreserved at - 80°C until further full sample processing.

### Total-seq C antibody staining and 10x Chromium loading

#### Newcastle

200,000 cells from each donor were stained with Human TruStain FcX™ Fc Blocking Reagent (Biolegend 422302) for 10 min at room temperature. The cells were then stained with the custom panel Total-seq C (Biolegend 99813; see **Supplementary Table 2**) for 30 min at 4°C. Cells were then washed twice with PBS supplemented with 2% (v/v) FCS and 2 mM EDTA (Sigma) before resuspending in PBS and counting. 20,000-30,000 cells per sample were loaded onto the 10x Chromium controller using Chromium NextGEM Single Cell V(D)J Reagent kits v1.1 with Feature Barcoding technology for Cell Surface Protein (10x Genomics) according to the manufacturer’s protocol.

#### Cambridge

Half a million viable cells were resuspended in 25 μL of FACS buffer and incubated with 2.5 μL of Human TruStain FcX™ Fc Blocking Reagent (BioLegend 422302) for 10 min at 4°C. The TotalSeq-C™ antibody cocktail (BioLegend 99813; see **Supplementary Table 2**) was centrifuged at 14,000 g at 4°C for 1 min, resuspended in 52 μL of FACS buffer, incubated at room temperature for 5 min and centrifuged at 14,000 g at 4°C for 10 min. 25 μL were subsequently added to each sample pool and incubated for 30 min at 4°C in the dark. Pools were washed 3 times with 27 volumes (1.4 mL) of FACS buffer, followed by centrifugation at 500 g for 5 min. The pellet was resuspended in 62.5 µL of 1 x PBS + 0.04% BSA (Ambion, #AM2616), filtered through a 40 μm cell strainer (Flowmi, H13680-0040) and viable cells of each sample pool were counted in an improved Neubauer chamber using Trypan blue. 50,000 live cells (up to a maximum of 60,000 total cells) for each pool were processed using Single Cell V(D)J 5’ version 1.1 (1000020) together with Single Cell 5’ Feature Barcode library kit (1000080), Single Cell V(D)J Enrichment Kit, Human B Cells (1000016) and Single Cell V(D)J Enrichment Kit, Human T Cells (1000005) (10x Genomics) according to the manufacturer’s protocols.

#### UCL/Sanger

Frozen PBMC samples were thawed quickly in a water bath at 37°C. Warm RPMI1640 medium (20-30 mL) containing 10% FBS was added slowly to the cells before centrifuging at 300 g for 5 min, the pellet was then washed with 5 mL RPMI1640-FBS and centrifuged again (300 g for 5 min). The PBMC pellet was collected and cell number and viability determined using Trypan blue. PBMCs from four different donors were then pooled together at equal numbers (1.25×10^5^ PBMCs from each donor) to make up 5.0×10^5^ cells in total. The remaining cells were used for DNA extraction (Qiagen, 69504). The pooled PBMCs were stained with TotalSeq-C antibodies (Biolegend, 99814) according to manufacturer’s instructions. After incubating with 0.5 vial of TotalSeq-C for 30 min at 4°C, PBMCs were washed three times by centrifugation at 500 g for 5 min at 4°C. PBMCs were counted again and processed immediately for 10x 5’ single cell capture (Chromium Next GEM Single Cell V(D)J Reagent Kit v1.1 with Feature Barcoding technology for cell Surface Protein-Rev D protocol). Two lanes of 25,000 cells were loaded per pool on a 10x chip.

### Library preparation and sequencing

#### Newcastle and UCL/Sanger

Gene expression, TCR enriched and BCR enriched libraries were prepared for each sample according to the manufacturer’s protocol (10x Genomics). Cell surface protein libraries were subjected to double the manufacturer’s recommended primer concentration and 7-8 amplification cycles during the sample index PCR to reduce the likelihood of daisy chains forming. Libraries were pooled per patient using the following ratio 6:2:1:1 for gene expression, cell surface protein, TCR enriched and BCR enriched libraries. All libraries were sequenced using a NovaSeq 6000 (Illumina) to achieve a minimum of 50,000 paired end reads per cell for gene expression and 20,000 paired end reads per cell for cell surface protein, TCR enriched and BCR enriched.

#### Cambridge

The samples were subjected to 12 cycles of cDNA amplification and 8 cycles for the cell surface protein library construction. Following this, the libraries were processed according to the manufacturer’s protocol. Libraries were pooled per sample using a ratio 9:2.4:1:0.6 for gene expression, cell surface, TCR enriched and BCR enriched libraries. Samples were sequenced using a NovaSeq 6000 (Illumina), using S1 flowcells.

### Alignment and quantification

Droplet libraries were processed using Cellranger v4.0. Reads were aligned to the GRCh38 human genome concatenated to the SARS-Cov-2 genome (NCBI SARS-CoV-2 isolate Wuhan-Hu-1) using STAR^55^ and unique molecular identifiers (UMIs) deduplicated. CITE-seq UMIs were counted for GEX and ADT libraries simultaneously to generate feature X droplet UMI count matrices.

### Doublet identification

#### Newcastle

*Scrublet* (v0.2.1) was applied to each sample to generate a doublet score. These formed a bimodal distribution so the tool’s automatic threshold was applied.

#### Cambridge

Non-empty droplets were called within each multiplexed pool of donors using the *emptyDrops* function implemented in the Bioconductor package *DropletUtils*, using a UMI threshold of 100 and FDR of 1%. The probability of being a doublet was estimated for each cell per sample (that is one 10x lane) using the “*doubletCells*” function in *scran* based on highly variable genes (HVGs). Next, we used “*cluster_walktrap*” on the SNN-Graph that was computed on HVGs to form highly resolved clusters per sample. Per-sample clusters with either a median doublet score greater than the median + 2.5 x MAD or clusters containing more than the median + 2.5 MAD genotype doublets were tagged as doublets. This was followed by a second round of highly-resolved clustering across the whole data set, in which again cells belonging to clusters with a high proportion (> 60%) of cells previously labelled as doublets were also defined as doublets.

#### UCL/Sanger

For pooled donor CITE-seq samples, the donor ID of each cell was determined by genotype-based demultiplexing using *souporcell* version 2^56^. *Souporcell* analyses were performed with ‘skip_remap’ enabled and a set of known donor genotypes given under the ‘common_variants’ parameter. The donor ID of each souporcell genotype cluster was annotated by comparing each souporcell genotype to the set of known genotypes. Droplets that contained more than one genotype according to souporcell were flagged as ‘ground-truth’ doublets for heterotypic doublet identification. Ground-truth doublets were used by *DoubletFinder* 2.0.3^57^ to empirically determine an optimal ‘pK’ value for doublet detection. *DoubletFinder* analysis was performed on each sample separately using 10 principal components, a ‘pN’ value of 0.25, and the ‘nExp’ parameter estimated from the fraction of ground-truth doublets and the number of pooled donors.

### CITE-seq background signal removal

Background antibody- and non-specific staining was subtracted from ADT counts in each data set from the 3 data acquisition sites separately. ADT counts for each protein were first normalised using counts per million (CPM) and log transformed, with a +1 pseudocount. To estimate the background signal for each protein, a 2-component gaussian mixture model (GMM), implemented in the *mclust* R package function *Mclust*, was fit across the droplets with a total UMI count > 10 and < 100 from each experimental sample separately. The mean of the first GMM component for each protein was then subtracted from the log CPM from the QC-passed droplets in the respective experimental sample.

### Quality control, normalisation, embedding and clustering

Combined raw data from the three centres was filtered to remove those that expressed fewer than 200 genes and >10% mitochondrial reads. Data was normalised (scanpy: *normalize_total*), log+1 corrected (scanpy: *log1p*) and highly variable genes identified using the Seurat vst algorithm (scanpy: *highly_variable_genes*). Harmony was used to adjust principal components by sample ID and used to generate the neighbourhood graph and embedded using UMAP. Clustering was performed using the Leiden algorithm with an initial resolution of 3. For initial clustering, differentially expressed genes were calculated using Wilcoxon rank-sum test.

### Cluster differential abundance testing

Numbers of cells of each cell subtype were quantified in each patient and control sample (donors) to compute a cell type X donor counts matrix. Cell type abundance counts were modelled as a function of either disease severity or days from symptom onset, adjusting for age, gender and batch, in a NB GLM, implemented in the Bioconductor package *edgeR*. Counts were normalised in the model using the (log) of the total numbers of all cells captured for each donor. Hypothesis testing was performed using quasi-likelihood F-test for either a linear or quadratic trend across disease severity groups (asymptomatic > mild > moderate > severe > critical), or comparing healthy controls to SARS-CoV-2 infected donors (healthy vs. all asymptomatic, mild, moderate, severe & critical). Differentially abundant cell types were determined using a 10% false discovery rate (FDR). Due to compositional differences across sites, when analysing differential abundance of myeloid populations (figure 2), only samples from Ncl and UCL/Sanger were included.

### Relative importance of metadata on cell type composition

The number of cells for each sample (*N*=110 samples in total with complete metadata) and cell type (18 different cell types in total) combination was modelled with a generalised linear mixed model with a Poisson outcome. The 5 clinical factors (COVID-19 swab result, age, sex, disease severity at day 0 and days from onset) and the 2 technical factors (patient and sequencing centre) were fitted as random effects to overcome the collinearlity among the factors. The effect of each clinical/technical factor on cell type composition was estimated by the interaction term with the cell type (see **Supplementary Note 1** for detail). The likelihood ratio test was performed to assess the statistical significance of each factor on cell type abundance by removing one interaction term from the full model at a time. The number of factors was used to adjust multiple testing with the Bonferroni approach. The ‘glmer’ function in the *lme4* package implemented on R was used to fit the model. The standard error of variance parameter for each factor was estimated using the *numDeriv* package.

### Cydar Analysis

We utilized cydar to identify changes in cell composition across the different severity groups based on the protein data alone. First, the background-corrected protein counts from the three different sites were integrated using the ‘*fastMNN*’ method (k = 20, d = 50, cos.norm = TRUE) in *scran*^*58*^. The batch-corrected counts for 188 proteins (4 rat/mouse antibody isotypes were removed) were then used to construct hyperspheres using the ‘*countCells*’ function (downsample = 8) with the tolerance parameter chosen so that each hypersphere has at least 20 cells which was estimated using the ‘*neighborDistances*’ function. To assess whether the abundance of cells in each hypersphere are associated with disease status, hypersphere counts were analyzed using the quasi-likelihood (QL) method in *edgeR*. After filtering out hyperspheres with an average count per sample below 5 we fitted a mean-dependent trend to the NB dispersion estimates. The trended dispersion for each hypersphere was used to fit a NB GLM using the log-transformed total number of cells as the offset for each sample and blocking for sex, age and batch. The QL F-test was used to compute *P* values for each hypersphere which were corrected for multiple testing using the spatial FDR method in *cydar*.

### Comparisons of PBMC annotation using the Azimuth tool

The final annotation of PBMCs was compared to a published PBMC annotation using the *Azimuth* tool (http://azimuth.satijalab.org/app/azimuth). Because of size restrictions of 100,000 cells, our data was subsampled to 10% of the total cells. After running the algorithm, results with a prediction score < 0.5 were removed (5.8% of total removed). For each cluster in the COVID-19 PBMC data, the percentage of cells mapped to each cluster in the *Azimuth* annotation was calculated.

### Interferon, TNF and JAK-STAT response scoring

A list of genes related to response to type I interferons was obtained from the GSEA Molecular Signatures Database (MSigDB) (GO: 0034340). Enrichment of the interferon score was measured using the *tl*.*score_genes* tool in *scanpy* which subtracts the average expression of all genes in the dataset from the average expression of the genes in this list. The scores were averaged across clusters and clinical status and expressed as a fold-change over the interferon score in the equivalent healthy cluster.

### kBET analysis

The *kBET*^*59*^ algorithm (https://github.com/theislab/kBET) was run for each cluster defined in Fig. 1 using the Uniform manifold and projection (UMAP) coordinates generated from Harmony-adjusted principal components, and the sample number as the batch factor. The same procedure was then performed using the same annotation but using the UMAP coordinates generated from non-Harmony-adjusted principal components. The resultant rejection rates were averaged across clusters and compared using a Wilcoxon paired signed rank test.

### Bronchoalveolar lavage data analysis

ScRNAseq data from BAL was obtained from GEO (accession number GSE145926^29^). Raw data was analysed using the same pipeline as PBMC data, specifically using the same quality control cut-offs (min of 200 genes and <10% mitochondrial reads/cell) and batch-corrected using Harmony by donor ID. To gain greater resolution of mononuclear phagocytes the DC and macrophages were analysed with further rounds of sub-clustering to identify DC1, DC2 and mature DC.

### PAGA analysis of blood monocytes and BAL macrophages

Annotated raw expression datasets of BAL macrophages and COVID-19 PBMCs were merged and data log-normalised and scaled as for the original datasets. The top 3000 highly variable genes were chosen using the *Seurat* “vst” method and used for downstream analysis. Principal components were batch corrected by donor and used to build a neighborhood graph. The PAGA tool in *scanpy* (*tl*.*paga*) was used to generate the abstracted graph between clusters.

### CellphoneDB

*CellphoneDB*^*60*^ was used to assess putative interactions between monocytes (CD14_mono, CD83_CD14_mono, C1_CD16_mono, CD16_mono, Prolif_mono) and platelets. The tool was run for 100 iterations and an expression threshold of 0.25 (limiting the analysis to genes expressed by 25% of cells). For downstream analysis we focused on interactions between platelets and any monocyte subset.

### HPSC commitment scoring

HPSCs were subsetted from the data and Leiden clusters generated using the same pipeline and parameters as for the whole PBMC dataset. Differentially expressed genes between the HSPC clusters that showed evidence of lineage commitment (MK, Erythroid and Myeloid) were calculated using *FindAllMarkers* tool in *Seurat* (with thresholds of genes expressed by 25% of cells and with a log fold-change of 0.25) and genes with an adjusted p-value cut-off of 0.05 were used to generate gene signatures for each. Enrichment of these signatures in the CD38 negative and CD38 positive HSPC clusters were calculated using the *tl*.*score_genes* in *scanpy*. The average expression of these enrichment scores in the CD38 negative and CD38 positive HSPC clusters was calculated and normalised to their expression in healthy patients.

### Multiplex cytokine analysis

Serum was obtained from peripheral blood in red topped serum Vacutainers® (BD, 367815) and allowed to clot for at least 30 min before centrifugation (800 g for 10 min) to separate the serum. After collection, serum was frozen at −80°C and thawed on ice on the day of experiment. The assay was carried out using the Cytokine/Chemokine/Growth Factor 45-Plex Human ProcartaPlex™ Panel 1 kit (Invitrogen, EPX450-12171-901), utilising the Luminex xMAP technology and according to the manufacturer’s protocol. Each sample was run in duplicate. The values of each analyte were detected using the MAGPIX® system and analysed using the ProcartaPlex Analyst version 1.0 Software (ThermoFisher Scientific).

### Re-stimulation of PBMC with SARS-Cov-2 peptide S

Purified PMBC were thawed at 37°C, transferred into a 15 mL tube with 10 mL pre-warmed complete culture media RPMI-1640 medium (Sigma Aldrich, R0883) supplemented with 10% (v/v) FCS (Gibco, 10270-106), 1% (v/v) Penicillin/Streptomycin (100 U/mL and 100 μg/mL respectively; Sigma Aldrich, P0781) and 1% (v/v) L-Glutamine (2 mM; Sigma Aldrich, G7513), referred as RPMI10, followed by centrifugation at 500 g for 5 min. Cell pellet was resuspended in 500 μL RPMI10 with added DNAse (1 μg/mL, Merck, 10104159001), divided into 5 wells of round bottom 96-well plate and left to rest at 37°C for an hour. Cells were stimulated with SARS-CoV-2 PepTivator peptide S for pan-HLA (2 μg/mL, Miltenyi Biotech, 136-126-700) and PMA/Ionomycin as a control (2 μL/mL, Cell Activation cocktail, Biolegend, 423301), and incubated at 37°C for 2 h. Negative controls were left untreated. Brefeldin A (2 μg/mL, GolgiPlug, BD Bioscience, 555029) and anti-CD107a-BB700 antibody (1:50, clone H4A3, BD Bioscience, 566558) was added for additional 4 h into all conditions. Cells were stained for detection of activation induced markers and intracellular cytokines 6 h after stimulation and subjected to flow cytometry.

### Flow Cytometry of stimulated cells

PBMC stimulated for 6 h with the SARS-Cov-2 peptide were washed with PBS, and cell surface stained for 1 h at room temperature: anti-CD14-FITC (1:50, clone M5E2, BD Biosciences, 555397), anti-CD19-FITC (1:50, clone 4G7, BD Biosciences, 345776), anti-CD137-Pe-Dazzle594 (1:50, clone 4B4-1, Biolegend, 309826), anti-CCR7-PE-Cy7 (1:50, clone G043H7, Biolegend, 353226), anti-CD45RO-APC-H7 (1:50, clone UCHL1, BD Biosciences, 561137), anti-CD28-BV480 (1:50, clone CD28.2, BD Biosciences, 566110), anti-CD4-BV785 (1:100, clone SK3, Biolegend, 344642), anti-CD3-BUV395 (1:50, clone UCHT1, BD Biosciences, 563546), anti-CD8-BUV496 (1:100, clone RPA-T8, BD Biosciences, 564804), anti-CD25-BUV737 (1:100, clone 2A3, BD Biosciences, 612806) and viability dye Zombie Yellow (1:200, Biolegend, 423104). Cells were washed with PBS 2% (v/v) FCS, fixed with 4% (w/v) paraformaldehyde (ThermoFisher Scientific, 28908) and kept at 4°C overnight. Subsequently, cells were washed with PBS, permeabilized with Perm/Wash buffer (BD Biosciences, 554723) according manufacturer’s instruction, and stained with intracellular antibodies for 1 h on ice: anti-IL10-PE (1:10, clone JES3-19F1, BD Biosciences, 559330), anti-IFN-αPC (1:25, Miltenyi Biotec, 130-090-762), anti-TNF-AF700 (1:50, clone MAb11, Biolegend, 502928), anti-IL2-BV421 (1:100, clone 5344.111, BD Biosciences, 562914), anti-CD154-BV605 (1:50, clone 24-31, Biolegend, 310826). Cells were washed, transferred to flow cytometry 5 mL tubes, and acquired on Symphony A5 flow cytometer (BD Biosciences). Data were analysed by FlowJo V10 (BD Biosciences).

### GSEA analysis

Pre-ranked gene set analysis (prGSEA) on MSigDB v7.2 Hallmark genesets^61^ was performed using pre-ranked gene lists with fgsea^62^ in R. Genes were pre-ranked according to signed -log_10_ *P*-values for all prGSEA procedures. For B cells, generation of rank gene list was performed using Wilcoxon rank sum test (via *tl*.*rank_genes_groups* in *scanpy*) with each Day 0 COVID statuses (asymptomatic to symptomatic critical) as the “tests” versus Day 0 Healthy samples as “reference/control”.

### T cell clustering, annotation and visualisation

Droplets labelled as T cells (“CD4”, “CD8”, “Treg”, “MAIT”, “gdT”) were subset from those in Fig. 1B and re-clustered using a set of HVGs calculated within each batch, the union of which were used to estimate the first 50 principal components across cells using the *irbla* R package. Batch effects were removed across the first 30 PCs using the *fastMNN*^*58*^ implementation in the Bioconductor package *batchelor* (k=50). A k-nearest neighbour graph (k=20) was computed across these 30 batch-integrated PCs using the *buildKNNGraph* function implemented in the Bioconductor package *scran*, which was then used to group cells into connected communities using Louvain^63^ clustering implemented in the R package *igraph*. Clusters that displayed mixed profiles of T and other lymphoid lineages, i.e. CD19, CD20 and immunoglobulin genes, were classed as doublets and removed from down-stream analyses. Clusters indicative of NK cells (CD3^-^CD56^+^) were subsequently annotated as such and removed from T cell analyses. Remaining clusters were annotated using a combination of canonical protein & mRNA (italicised) markers for major αβ T cells (CD4, CD8, CCR7, CD45RA, CD45RO, CD62L, CD27, CD38, CD44, CXCR5, CD40LG *CCR7, FOXP3, IKZF2*), γδT cells (Vγ9, Vγ2, *TRGV9, TRDV2*) and invariant T cells; MAIT (Vα24-Jα18, *TRAV1*.*2*), NKT (CD3, CD16, CD56, *NCAM1, NCR1, FCGR3A*). Polarized CD4^+^ T cell annotations were refined using the combination of transcription factor genes and expressed cytokines for the respective helper T cell types: Th1 (*IFNG, TBX21, TNFA*), Th2 (*GATA3, IL4, IL5*), Th17 (*RORC, IL17A, IL17F, IL21*). Where clusters appeared heterogeneous in their expression of T cell lineage markers, single cell annotations were refined based on the co-expression of specific marker gene and protein pairs. Dot plots to visualise marker protein and mRNA expression across clusters were generated using the R package *ggplot2*. UMAP^64^ was used to project all single T cells into a 2D space (k=31) using the first 30 batch-integrated PCs as input using the R package *umap*.

### T cell differential gene expression analysis

Differential gene expression (DGE) analysis was performed across COVID-19 disease severity groups, ordered from healthy > asymptomatic > mild > moderate > severe > critical. Donor pseudo-bulk samples were first created by aggregating gene counts for each annotated T cell type, within each donor, where there were at least 20 cells of that type. Genes with fewer than 3 counts in any given pseudo-bulk, or fewer than 5 counts in total across donor pseudo-bulk samples, were removed prior to analysis. DGE testing was performed using a negative binomial generalized linear model (NB GLM) implemented in the Bioconductor package *edgeR*^*65,66*^. *Statistically significant DE genes were defined with FDR < 0*.*1. Functional annotation enrichment was performed using the Bioconductor package enrichR*^*67*^. Up- and down-regulated DE genes in each T cell type were used as input, testing separately against the MSigDB Hallmark 2020 and Transcription Factor Protein-Protein Interactions gene sets. Significant enrichments were defined with 1% FDR.

### T cell receptor analysis

Single-cell TCRs were computed from the TCR-seq data using Cellranger v4.0.0. The unfiltered output of reconstructed TCR contigs across all 3 sites (Newcastle, Cambridge, UCL) were combined prior to filtering using: 1) full length CDR3, 2) droplet barcode matched a T cell droplet, 3) productive CDR3 spanning V+J genes. Chain-specific TCR clones were defined for each observed α and β chain by first concatenating the V, J and identical CDR3 nucleotide sequences. For each single T cell, these chains were then combined to form a single clonotype, removing cells that contained: 1) > 2 β chains and > 2 α chains, 2) a single α or a single β chain only. T cells with exactly 2 β chains and 1 α chain, or those with exactly 2 α chains and 1 β chain were retained. TCR clonotypes were counted within each donor sample, and expanded clones were defined where > 1 cell was assigned to the TCR clonotype.

The proportion of expanded clones as a function of a linear trend across disease severity groups was modelled using logistic regression, adjusted for age, gender and batch. A separate model was run for each T cell subtype which contained at least 5 cells assigned to the expanded TCR clonotypes. Linear trend p-values were corrected for multiple testing using the Benjamini & Hochberg procedure^68^.

The TE:EM ratio was calculated within each donor, using the number of observed expanded clonotypes. The TE:EM ratio change across COVID-19 severity was tested using a robust linear model implemented in the R package *robustbase*, regressing TE:EM ratio on disease severity as an ordered linear variable (asymptomatic > mild > moderate > severe > critical), adjusted for age, gender and batch. Statistical significance was defined based on the linear trend across disease severity (p ≤ 0.01).

### Differential correlation analysis

Changes in the correlations between PBMC cell types were computed using a differential correlation analysis, implemented in the R package *DCARS*^*69*^. Cell type proportions were computed by normalizing the counts of each cell type within each donor by the total number of cells captured for that donor sample. Donor samples were ranked according to their disease severity (healthy > asymptomatic > mild > moderate > severe > critical). Differential correlation analysis was then performed between CD4.Tfh vs all B cell types. Statistically significant differentially correlated cell types were defined with empirical p-value ≤ 0.05, estimated from 10,000 permutations.

### BCR V(D)J analysis

Single-cell V(D)J data from the 5’ Chromium 10x kit were initially processed with *cellranger-vdj* (4.0.0). BCR contigs contained in *filtered_contigs*.*fasta* and *filtered_contig_annotations*.*csv* from all three sites were then pre-processed using *immcantion* inspired preprocessing pipeline^70^ implemented in the *dandelion* python package; *dandelion* is a novel single cell BCR-seq analysis package for 10x Chromium 5’ data. All steps outlined below are performed using *dandelion* v0.0.26 and is available at https://github.com/zktuong/dandelion.

### BCR preprocessing

Individual BCR contigs were re-annotated with *igblastn* v1.1.15 using the IMGT reference database (date downloaded: 30-June-2020)^71^ by calling *changeo*’s *AssignGenes*.*py* script and re-annotated contigs in *blast* format were parsed into the Adaptive Immune Receptor Repertoire (AIRR) standards 1.3 format with *changeo*’s *MakeDB*.*py* script. Amino acid sequence alignment information not present in the output from *blast* format were retrieved from re-annotation with *igblastn* in *airr* format. Heavy chain V-gene alleles were corrected for individual genotypes with TIgGER^72^ (v1.0.0) using a modified *tigger-genotype*.*R* script from *immcantation* suite. Germline sequences were reconstructed based on the genotype corrected V-gene assignments using *changeo*’s (v1.0.1) *CreateGermines*.*py* script; contigs which fail germline sequence reconstruction were removed from further analysis. Constant genes were re-annotated using blastn (v2.10.0+) with CH1 regions of constant gene sequences from IMGT followed by pairwise alignment against curated sequences to correct assignment errors due to insufficient length of constant regions.

### BCR filtering

Contigs assigned to cells that passed quality control on the transcriptome data were retained for further quality control assessment, which includes checks for: i) contigs with mismatched locus, V-, J- and constant gene assignments were removed from the analysis; ii) cell barcodes with multiple heavy chain contigs were flagged for filtering. Exceptions to this would be when a) the multiple heavy chain contigs were assessed to have identical V(D)J sequences but assigned as different contigs belonging to the same cell by *cellranger-vdj*, b) when there is a clear dominance (assessed by difference in UMI count) by a particular contig, and c) if and when there is presence of one IgM and one IgD contig assigned to a single cell barcode. In the first two cases, the contig with the highest UMI count is retained; iii) cell barcodes with multiple light chain contigs were flagged for filtering; iv) in situations where cell barcodes are matched with only light chain contigs, the contigs would be dropped from the V(D)J data but transcriptome barcode will be retained.

### B cell clone/clonotype definition

BCRs were grouped into clones/clonotypes based on the following sequential criterion that applies to both heavy chain and light chain contigs – i) identical V- and J-gene usage, ii) identical junctional CDR3 amino acid length, and iii) at least 85% amino acid sequence similarity at the CDR3 junction (based on hamming distance). Light chain pairing is performed using the same criterion within each heavy chain clone. Only samples collected at day 0 of the study were analyzed from this step onwards and clones/clonotypes were called across the entire dataset; the sample from one of the donors who was subsequently found to have a B cell malignancy was separated from the analysis and processed independently.

### B cell clone/clonotype network

Single-cell BCR networks were constructed using adjacency matrices computed from pairwise Levenshtein distance of the full amino acid sequence alignment for BCR(s) contained in every pair of cells within each disease severity cohort. Construction of the Levenshtein distance matrices were performed separately for heavy chain and light chain contigs and the sum of the total edit distance across all layers/matrices was used as the final adjacency matrix. To construct the BCR neighborhood graph, a minimum spanning tree was constructed on the adjacency matrix for each clone/clonotype, creating a simple graph with edges indicating the shortest edit distance between a B cell and its nearest neighbor. Cells with identical BCRs i.e. cells with a total pairwise edit distance of zero are then connected to the graph to recover edges trimmed off during the minimum spanning tree construction step. Fruchterman-reingold graph layout was generated using a modified method to prevent singletons from flying out to infinity in *networkx* (v2.5). Visualisation of the resulting single-cell BCR network is achieved via transferring the graph to relevant *anndata* slots, allowing for access to plotting tools in *scanpy*.

The use of the BCR network properties for computing gini indices was inspired from bulk BCR-seq network analysis methods where distribution of clone sizes and vertex sizes (sum of identical BCR reads) in BCR clone networks were used to infer the relationships between BCR clonality, somatic hypermutation and diversity^73^. However, there are challenges with native implementation of this approach for single-cell data. Firstly, to enable calculation of network-based clone/cluster and vertex/node size distribution, BCR networks needed to be reduced such that nodes/cells with identical BCRs had to be merged and counted; this required the re-construction of BCR networks per sample and discarding single-cell level information. Furthermore, the process of node contraction and counting of merging events requires significant computation time and resource. Secondly, this approach is dependent on sufficient coverage of the BCR repertoire, as the BCRs from the number of cells sampled (post-QC) may not necessarily recapitulate the entire repertoire, which may under- or over-represent merged counts for gini index calculation. We propose the use of node closeness centrality computed on each expanded clone (clone size > 1) as an alternative metric to emulate the statistics to adapt to the single-cell nature of the data; closeness centrality defines how close and central each node is with respect to other nodes in the graph and therefore cells with identical BCRs will have high closeness centrality scores, due to the way the BCR network is constructed in *dandelion*. Thus, we can quickly calculate if cells across clones, and/or samples overall, in the entire graph display proportionately/disproportionately high or low closeness centrality scores. One caveat to the current implementation is that it is only meaningful if there are clonotypes with at least two cells as scores will only be computed for non-singleton components of the graph. Gini indices are computed using *skbio*.*diversity*.*alpha*.*gini_index* (*scikit-bio* v0.5.6) with the *trapezoids* method after clone definition and network generation. Summary visualisation was performed using plotting tools in *seaborn* (v0.11.0).

### Definition of BCR convergence across patients

BCR overlap was determined by collapsing sharing incidence of V- and J-gene usage and CDR3 amino acid sequences, in both heavy and light chains, between individuals into a binarized format (1 or 0). The information is turned into an adjacency matrix where an edge is created between two individuals if there is at least one clonotype (at least 1 cell from each individual displays an identical combination of heavy and light chain V- and J-gene usage with allowance for somatic hypermutation at the CDR3 junctional region) that is similar between the two individuals. Visualisation is achieved using the *CircosPlot* function from *nxviz* package (v0.6.2).

