## Extended Data for "The cellular immune response to COVID-19 deciphered by single cell multi-omics across three UK centres"

### Extended Data 1

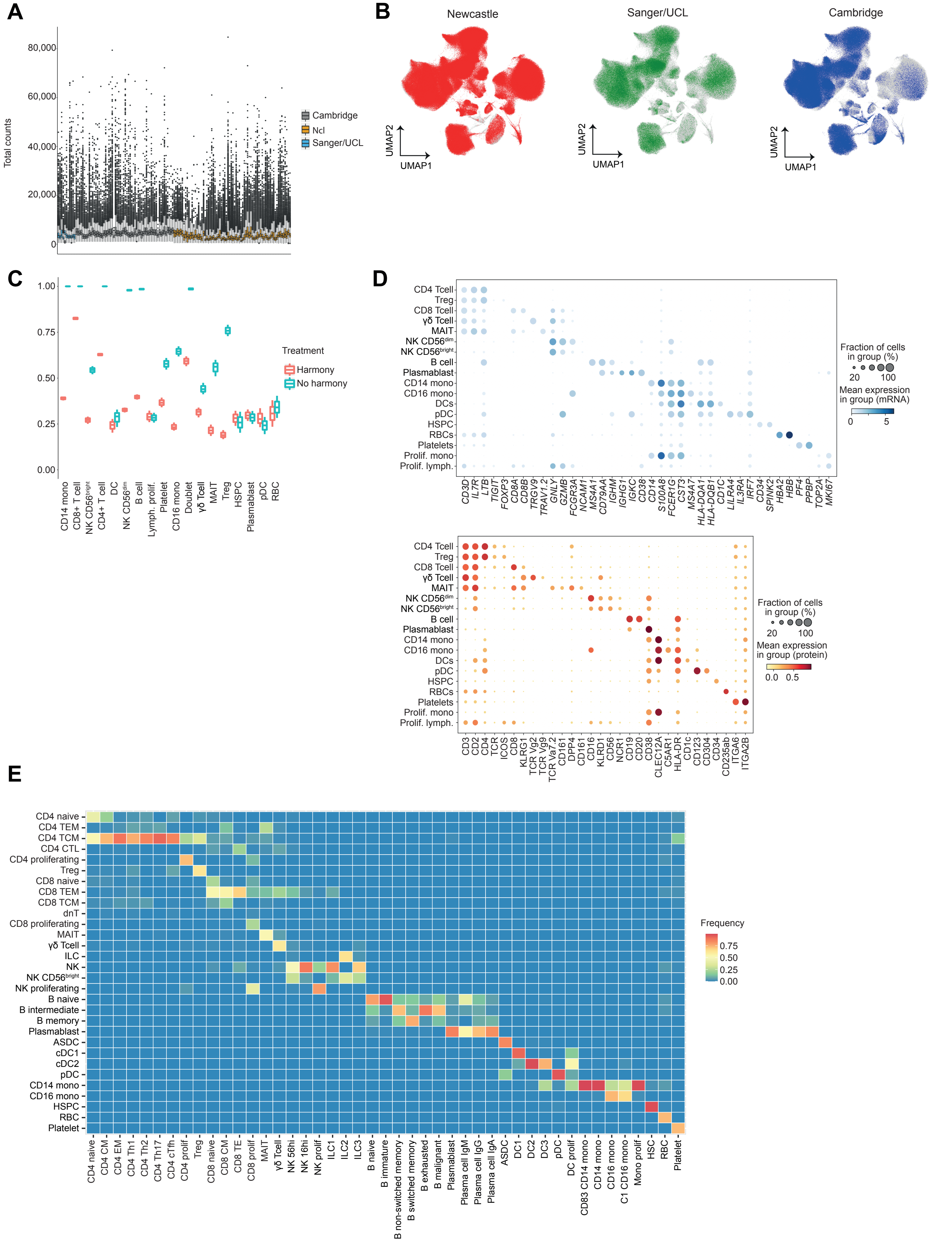

### Extended Data 2

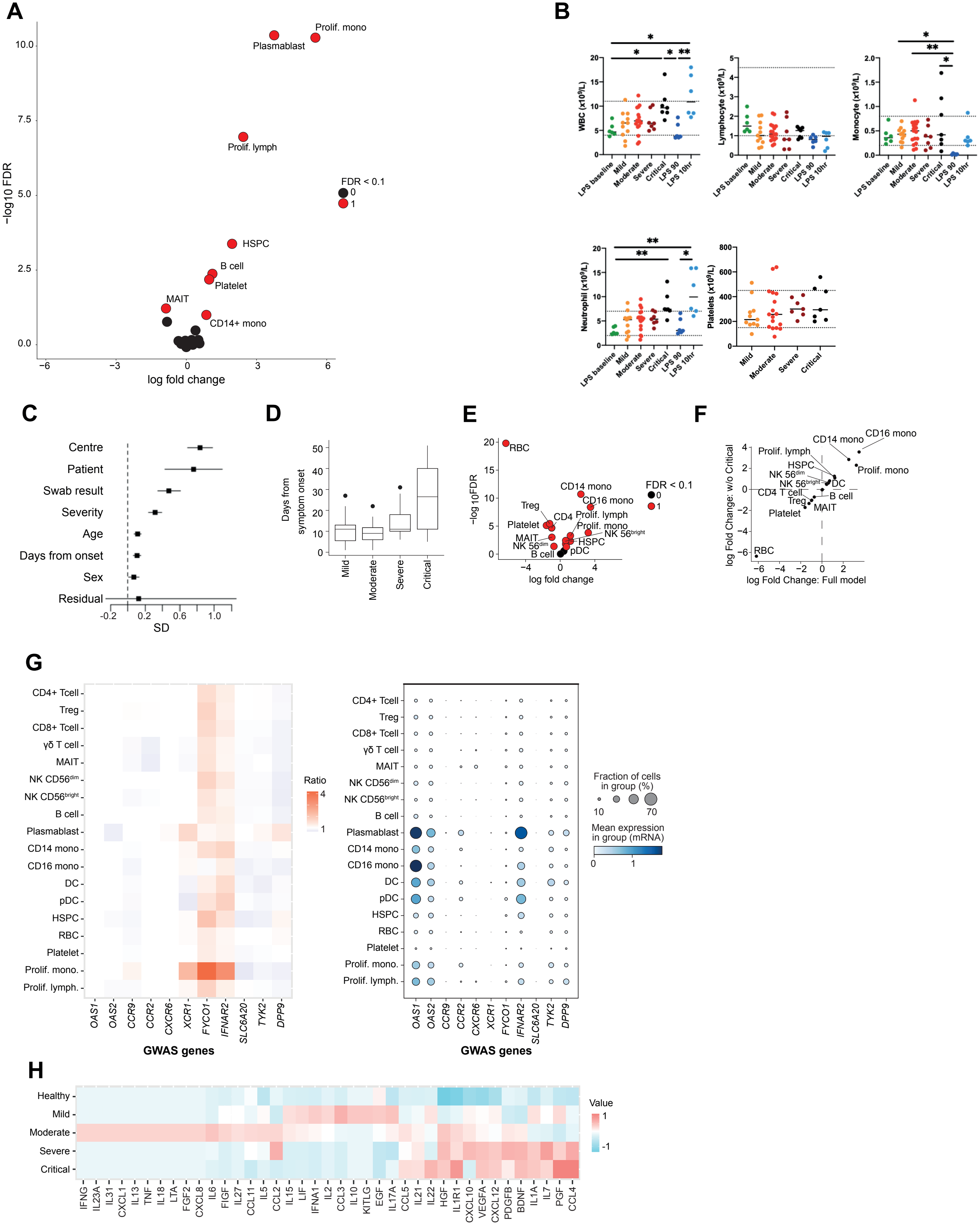

### Extended Data 3

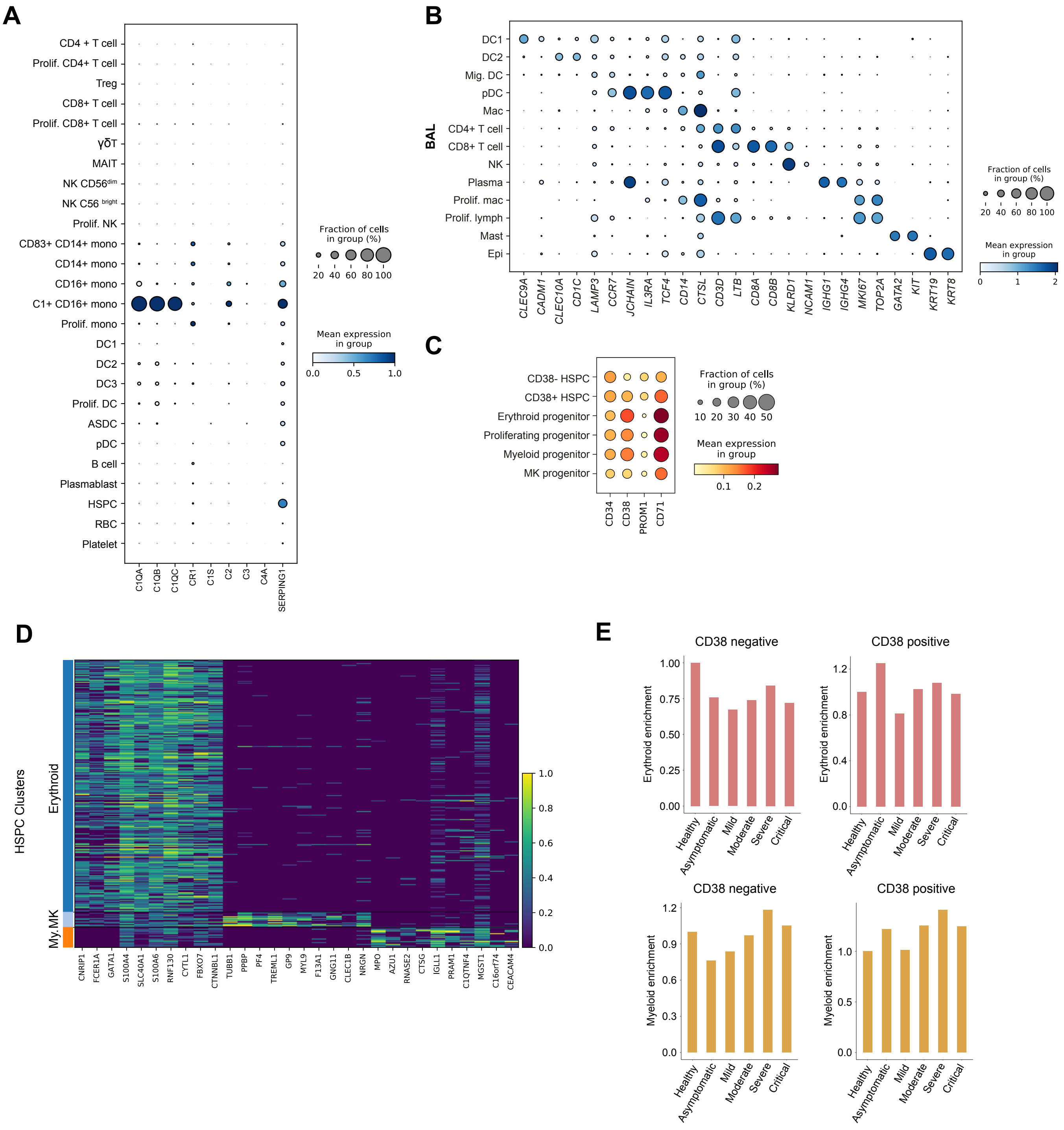

### Extended Data 4

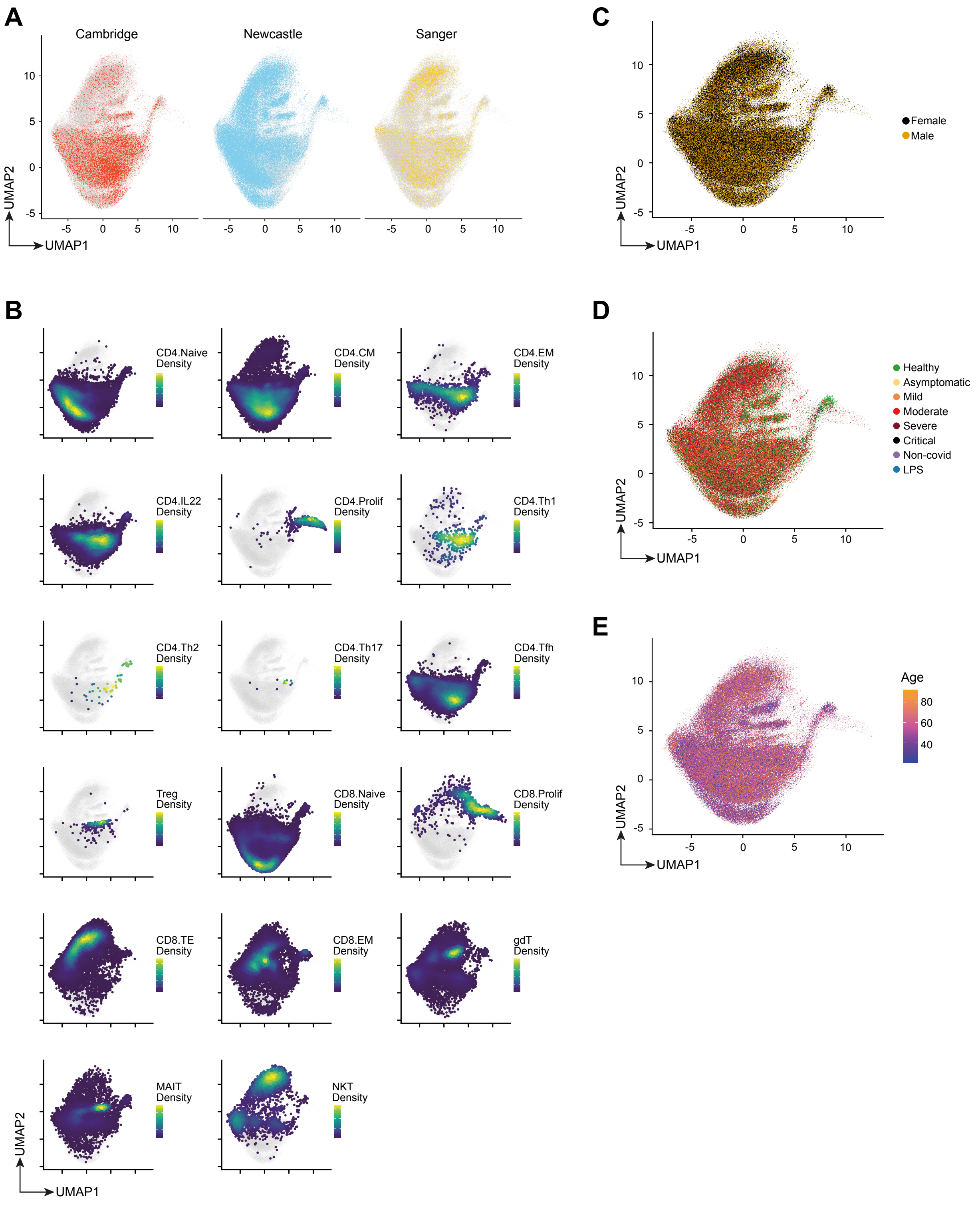

### Extended Data 5

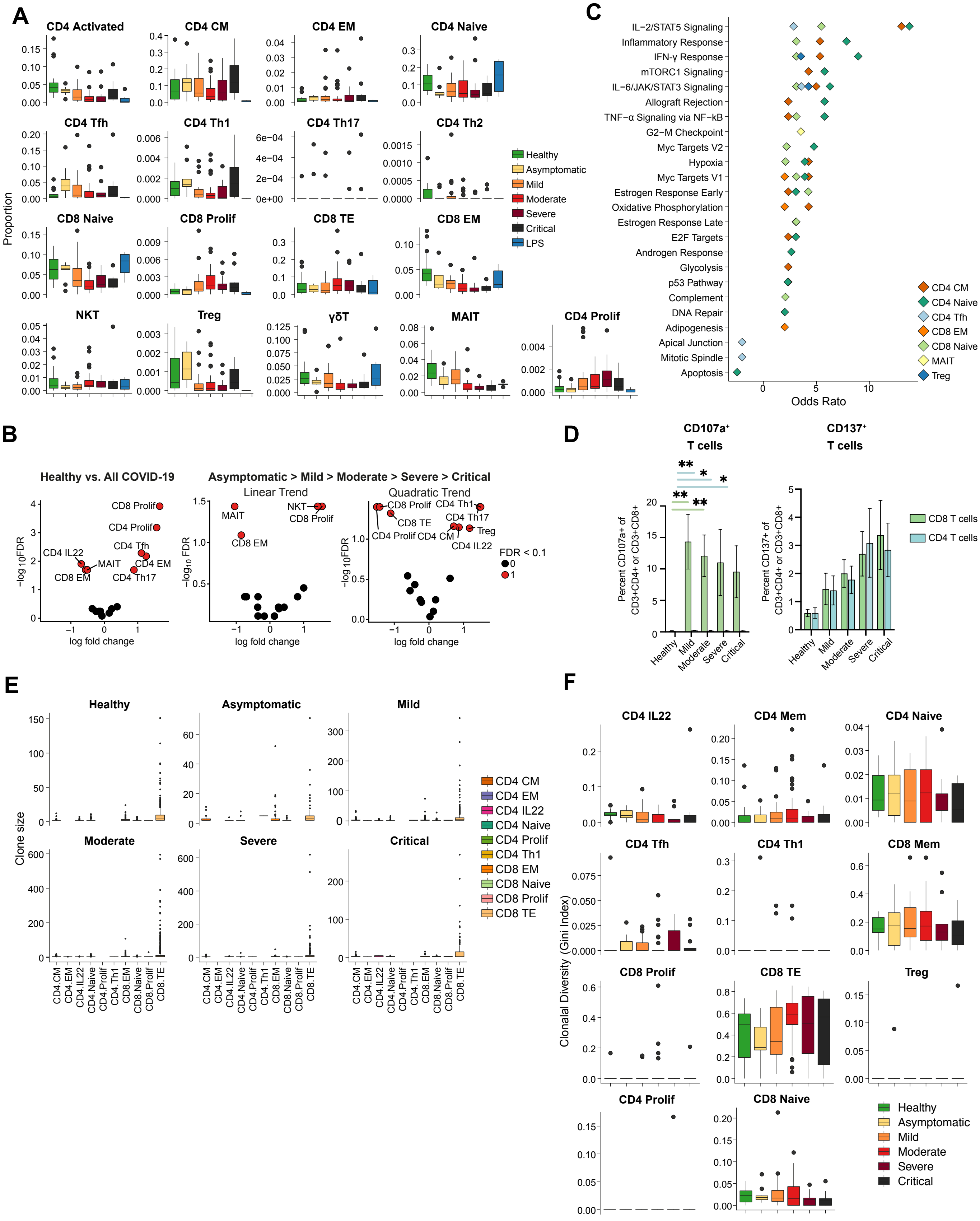

### Extended Data 6

**A**

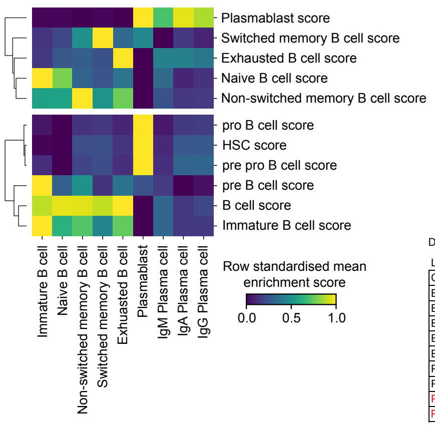


**B**

Kruskal-Wallis test

| | Plasmablast | Plasma cell | | | B_exhausted | B_memory | | B_naive | B_immature |
|---|---|---|---|---|---|---|---|---|---|
| | | IgM | IgA | IgG | | switched | non-switched | | |
| Healthy vs. Asymptomatic | | | | | | | | | |
| Healthy vs. Mild | ** | | | ** | | * | | | |
| Healthy vs. Moderate | *** | | * | ** | | * | | | * |
| Healthy vs. Severe | *** | | ** | ** | | | | | ** |
| Healthy vs. Critical | ** | | | * | | | | | |
| Healthy vs. Non_covid | | | | | | | | | |
| Healthy vs. LPS | | | | | | | | | |
| Asymptomatic vs. LPS | | | | | | | | | |
| Mild vs. LPS | | | | ** | | | | | |
| Moderate vs. LPS | * | | | ** | | | | | * |
| Severe vs. LPS | * | | * | ** | | | | | |
| Critical vs. LPS | | | | * | | | | | |
| Non_covid vs. LPS | | | | | | | | | |

Differential abundance test (asymptomatic > mild > moderate > severe > critical)

Linear changes

| CellType | logFC | logCPM | F | PValue | FDR |
|---|---|---|---|---|---|
| B_exhausted | 0.151 | 15.533 | 0.224 | 6.37E-01 | 8.20E-01 |
| B_immature | -0.138 | 16.203 | 0.268 | 6.06E-01 | 8.20E-01 |
| B_naive | -0.076 | 19.170 | 0.572 | 4.51E-01 | 8.12E-01 |
| B_non-switched_memory | 0.002 | 15.322 | 0.000 | 9.95E-01 | 9.95E-01 |
| B_switched_memory | -0.202 | 16.585 | 0.833 | 3.64E-01 | 8.12E-01 |
| Plasma_cell_IgA | 0.032 | 15.539 | 0.009 | 9.25E-01 | 9.95E-01 |
| Plasma_cell_IgG | 0.702 | 15.639 | 4.005 | 4.83E-02 | 1.45E-01 |
| Plasma_cell_IgM | 1.153 | 13.774 | 5.523 | 2.09E-02 | 9.40E-02 |
| Plasmablast | 1.145 | 15.784 | 7.526 | 7.31E-03 | 6.58E-02 |

Quadratic changes

| CellType | logFC | logCPM | F | PValue | FDR |
|---|---|---|---|---|---|
| B_exhausted | -0.046 | 15.533 | 0.025 | 8.74E-01 | 9.80E-01 |
| B_immature | 0.148 | 16.203 | 0.395 | 5.31E-01 | 6.83E-01 |
| B_naive | 0.147 | 19.170 | 2.744 | 1.01E-01 | 2.27E-01 |
| B_non-switched_memory | -0.006 | 15.322 | 0.001 | 9.80E-01 | 9.80E-01 |
| B_switched_memory | 0.155 | 16.585 | 0.613 | 4.36E-01 | 6.54E-01 |
| Plasma_cell_IgA | -0.402 | 15.539 | 1.663 | 2.00E-01 | 3.61E-01 |
| Plasma_cell_IgG | -1.016 | 15.639 | 9.926 | 2.20E-03 | 9.88E-03 |
| Plasma_cell_IgM | -2.071 | 13.774 | 24.085 | 3.95E-06 | 3.56E-05 |
| Plasmablast | -0.967 | 15.784 | 6.465 | 1.27E-02 | 3.80E-02 |

**C**

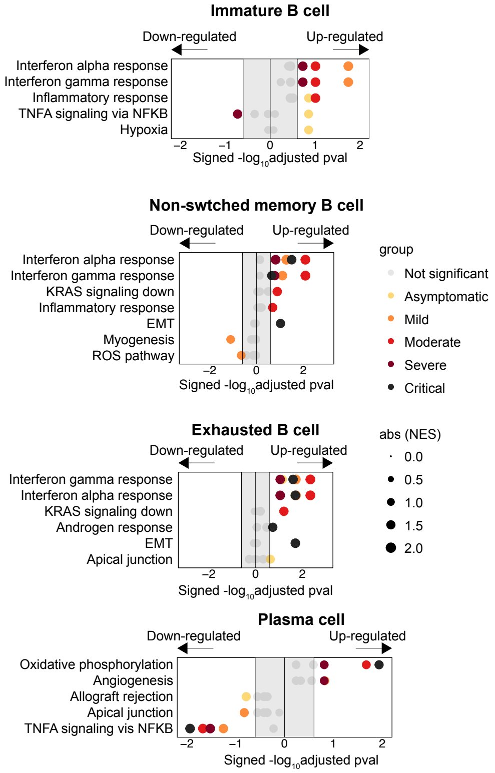


**D**

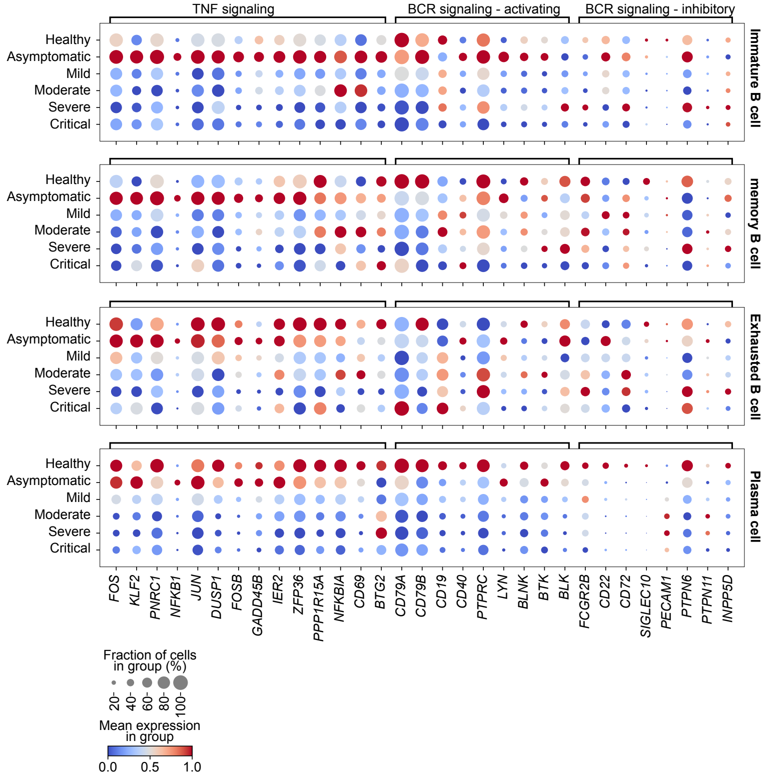

### Extended Data 7

A

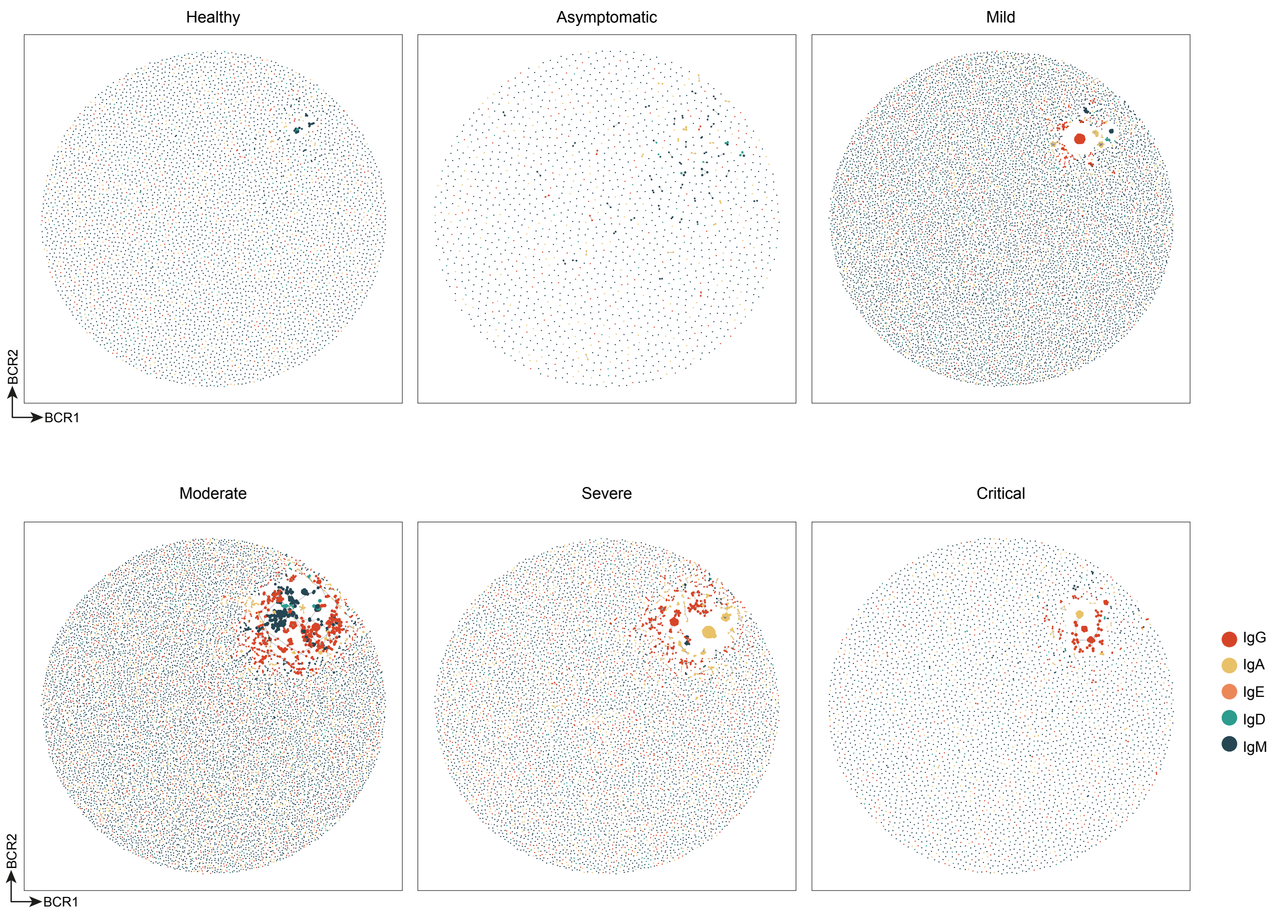


B

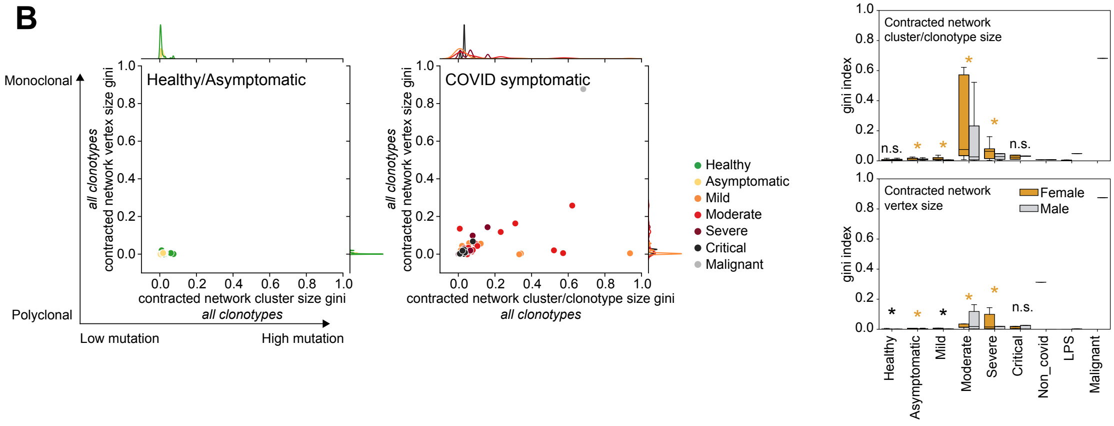

### Extended Data 8

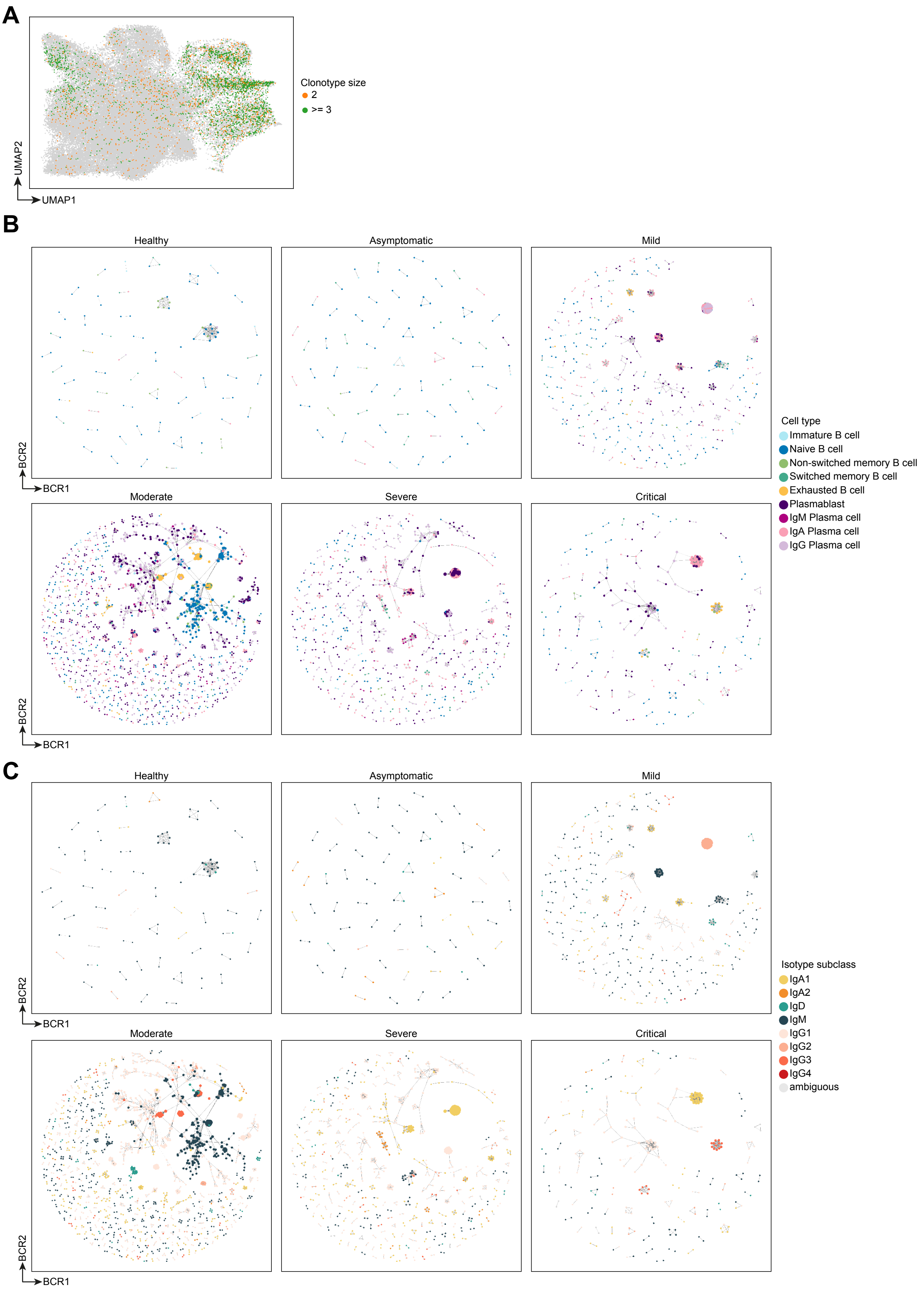
