## Supplementary Note 1 for "The cellular immune response to COVID-19 deciphered by single cell multi-omics across three UK centres"

### 1 Poission linear mixed model for cell type composition analysis

#### 1.1 Log linear model for two-way tables

Let $Y_{ij}$ be the cell type count observed from the sample $i$ ($i = 1, \ldots, N$) for the cell type $j$ ($j = 1, \ldots, J$). A simple test of independence between samples and cell types (to make sure there is no differential cell type abundance among samples) would be to fit a log-linear model for two-way tables [1]:

$$Y_{ij} \overset{i.i.d.}{\sim} \text{Pois}(\lambda_{ij}),$$
$$\log \lambda_{ij} = \mu + a_i + b_j + \varepsilon_{ij},$$

for $i = 1, \ldots, N$ and $j = 1, \ldots, J$. Here we assume $Y_{ij}$ follows a Poisson distribution with a mean $\lambda_{ij}$, the logarithm of which can be decomposed into the grand mean $\mu$, the sample mean $a_i$, the cell type mean $b_j$ and the interaction term $\varepsilon_{ij}$ (between sample $i$ and cell type $j$). In order to assess the two-way table is independent, we assume $\{a_i, b_j, \varepsilon_{ij}\}$ follow the independent normal distributions with variance parameters $\{\nu^2, \omega^2, \sigma^2\}$, such that

$$a_i \overset{i.i.d.}{\sim} \mathcal{N}(0, \nu^2), \quad b_j \overset{i.i.d.}{\sim} \mathcal{N}(0, \omega^2), \quad \varepsilon_{ij} \overset{i.i.d.}{\sim} \mathcal{N}(0, \sigma^2),$$

for $i = 1, \ldots, N$ and $j = 1, \ldots, J$, where the variance $\sigma^2$ is the parameter of interest. If there is no interaction (*i.e.*, no differential cell type abundance among samples), the variance estimate should become $\hat{\sigma}^2 \to 0$.

#### 1.2 Variance explained by sample metadata

Suppose $\sigma^2 > 0$, this model enables us to explore the relative importance of a wide range of clinical/technical factors in determining cell type composition. Let $x_{ik}$ be a value of the factor $k$ ($k = 1, \ldots, K$) for the sample $i$, which is either a numerical value (*e.g.* patient's age) or a categorical value of $L_k$ levels (*e.g.*, disease severity with $L_k = 6$: healthy, asymptomatic, mild, moderate, severe and critical). Then the mean of the poisson distribution can be extended with extra interaction terms between cell type and each of the $K$ factors, such that,

$$\log \lambda_{ij} = \mu + a_i + b_j + \sum_{k=1}^{K} \eta_{ijk} + \varepsilon_{ij}$$
$$\eta_{ijk} = \begin{cases} \boldsymbol{z}_{ik}^\top \boldsymbol{u}_{jk} & \text{factor } k \text{ is a categorical variable with } L_k \text{ levels,} \\ \tilde{x}_{ik} u_{jk} & \text{factor } k \text{ is a numerical variable } (L_k = 1), \end{cases}$$

where $\eta_{ijk}$ denotes the interaction effect between the cell type $j$ and the factor $k$ for the sample $i$, which is modelled by the interaction effect $\boldsymbol{u}_{jk} = (u_{jk1}, \ldots, u_{jkL_k})^\top$. Here $\tilde{x}_{ik}$ denotes the scaled value of $x_{ik}$ (*i.e.*, sample mean and variance of the numerical factor $k$ is 0 and 1) and $\boldsymbol{z}_{ik}^\top = (z_{ik1}, \ldots, z_{ikL_k})$ is a design vector whose element is

$$z_{jkl} = \begin{cases} 1 & x_{ik} = l, \\ 0 & \text{otherwise,} \end{cases}$$

for $l = 1, \ldots, L_k$. The interpretation of $u_{jkl}$ is the log fold change of the $j$th cell type abundance for the $l$th level of categorical factor $k$ against the grand mean. For a numerical factor $k$, $u_{jk}$ is a scalar value reflecting the log fold change of the $j$th cell type abundance in response to one unit change of scaled data $\tilde{x}_{ik}$.

Because the factors are colinear and often confounding each other (unless the study is the designed experiment), we further assume those interaction effects follow multivariate normal distributions:

$$\boldsymbol{u}_{jk} \overset{i.i.d.}{\sim} \mathcal{N}(\boldsymbol{\mu}_k, \delta_k^2 I_{L_k}),$$

where $\boldsymbol{\mu}_k$ denotes the mean vector around which the variance parameter $\delta_k^2$ is estimated, which reflects the relative contribution of each factor on cell type composition variation. Here the mean vector $\boldsymbol{\mu}_k$ is not the parameter of interest, therefore for the categorical factors, we regressed out from the model by assuming another multivariate normal distribution:

$$\boldsymbol{\mu}_k \overset{i.i.d.}{\sim} \mathcal{N}(0, \gamma_k^2 I_{L_k})$$

so that the number of parameters can be significantly reduced from $L_k$ to 1.

#### 1.3 Likelihood ratio test

To properly assess the statistical significance of each factor that explains a significant amount of interaction variation, we compared the the following two models:

$$H_0 : \delta_k^2 = 0$$
$$H_1 : \delta_k^2 > 0$$

Then the likelihood ratio test statistics follows the $\chi^2$ distribution with one degree of freedom under the null hypothesis ($H_0$). In order to adjust multiple testing, we used the number of factors (*i.e.,* $K$, which is the same as the number of variance parameters $\gamma_k^2$ for cell type interaction) for the total number of tests.

#### 1.4 Posterior mean and variance of random effects

In general, the generalised linear mixed model has no closed form of the marginal likelihood, because the integral with respect to random effects is intractable. Therefore an approximation becomes one of the natural alternatives. A well-known method of approximate integrals is named after Laplace (used in `lme4` package on `R`). Let $Y^\top = (Y_{11}, \ldots, Y_{NJ})$ be the vector of cell type counts and

$$\boldsymbol{u}^\top = (a_1, \ldots, a_N, b_1, \ldots, b_J, \boldsymbol{\mu}_1^\top, \ldots, \boldsymbol{\mu}_K^\top, \boldsymbol{u}_{11}^\top, \ldots, \boldsymbol{u}_{JK}^\top, \varepsilon_{11}, \ldots, \varepsilon_{NJ})$$

be the vector of all random effects, the marginal likelihood can be approximated as

$$p(Y) = \int p(Y|\boldsymbol{u})p(\boldsymbol{u})d\boldsymbol{u} \approx c|H|^{-\frac{1}{2}} \exp\{\mathcal{L}(\tilde{\boldsymbol{u}})\},$$

where $\mathcal{L}(\boldsymbol{u}) = \log p(Y|\boldsymbol{u})p(\boldsymbol{u})$ denotes the complete log likelihood function whose maximum is attained at $\boldsymbol{u} = \tilde{\boldsymbol{u}}$ with the first derivative $\mathcal{L}'(\tilde{\boldsymbol{u}}) = \mathbf{0}$ and the hessian matrix $H = -\mathcal{L}''(\tilde{\boldsymbol{u}})$, and $c$ denotes a constant multiplication. This gives an approximated posterior distribution of $\boldsymbol{u}$ given $Y$, such that

$$\boldsymbol{u}|Y \sim \mathcal{N}(\tilde{\boldsymbol{u}}, H^{-1}).$$

#### 1.5 Standard error of model parameters

The log marginal likelihood $\mathcal{L}(\boldsymbol{\theta}|Y) = \log p(Y)$ after integrating out the random effects $\boldsymbol{u}$ is a function of model parameters $\boldsymbol{\theta} = (\mu, \nu, \omega, \sigma, \gamma_1, \ldots, \gamma_K, \delta_1, \ldots, \delta_K)$. The standard error of $\boldsymbol{\theta}$ can be computed from the inverse matrix of the Fisher score matrix

$$\mathcal{I} = -\left.\frac{\partial^2 \mathcal{L}(\boldsymbol{\theta}|Y)}{\partial\boldsymbol{\theta}\partial\boldsymbol{\theta}^\top}\right|_{\boldsymbol{\theta}=\hat{\boldsymbol{\theta}}}$$

where the likelihood function attains its maximum value at $\boldsymbol{\theta} = \hat{\boldsymbol{\theta}}$ with $\mathcal{L}'(\hat{\boldsymbol{\theta}}|Y) = \mathbf{0}$.

##### 1.6 Overdispersion due to technical variation

Although the Poisson model does not explicitly take account of the overdispersion in the cell type count data (unlike Negative Binomial distributions), the interaction term $\varepsilon_{ij}$ between sample and cell type partly captures the discrepancy between $\mathbb{E}[Y_{ij}]$ and $\mathrm{Var}(Y_{ij})$, since

$$\mathrm{Var}(Y_{ij}|\boldsymbol{u}_{ij}) = \mathbb{E}[Y_{ij}|\boldsymbol{u}_{ij}] + \mathbb{E}[Y_{ij}|\boldsymbol{u}_{ij}]^2(e^{\sigma^2} - 1),$$

where $\boldsymbol{u}_{ij} = (a_i, b_j, \eta_{ij1}, \ldots, \eta_{ijK})^\top$. This fact suggests the model becomes overdispersed when $e^{\sigma^2} > 1$ given $\boldsymbol{u}_{ij}$.

### 2 R code

#### 2.1 Model fitting

We used the `lme4` package implemented on `R`. The log-linear model with Poisson outcome was implemented using `glmer` function with `family=poisson`, as follows:

```
library(lme4)
# Y : vector of counts whose length is N*J
full_model <- glmer(Y ~ (1|Celltype)
    + Age + (0+Age|Celltype)
    + Days_from_onset + (0+Days_from_onset|Celltype)
    + (1|Patient/Celltype)
    + (1|Sex/Celltype)
    + (1|Swab_result/Celltype)
    + (1|Status_D0/Celltype)
    + (1|Status_D3/Celltype)
    + (1|Status_D7/Celltype)
    + (1|Severity_D0/Celltype)
    + (1|Centre/Celltype)
    + (1|Sample/Celltype) # residual
  ,family=poisson
  ,data=metadata
  ,control=glmerControl(optimizer="bobyqa", optCtrl=list(maxfun=2e5)))
```

#### 2.2 Likelihood ratio test

In order to test whether a factor is meaningful, we simply dropped some of variance parameters from the model to compute likelihood ratio between the full model and the reduced model. For example, for the swap result:

```
# fitting reduced model without interaction between Swab_result and Celltype
reduced_model <- glmer(Y ~ (1|Celltype)
    + Age + (0+Age|Celltype)
    + Days_from_onset + (0+Days_from_onset|Celltype)
    + (1|Patient/Celltype)
    + (1|Sex/Celltype)
    + (1|Swab_result)
    + (1|Status_D0/Celltype)
    + (1|Status_D3/Celltype)
    + (1|Status_D7/Celltype)
    + (1|Severity_D0/Celltype)
    + (1|Centre/Celltype)
    + (1|Sample/Celltype) # residual
  ,family=poisson
  ,data=metadata
  ,control=glmerControl(optimizer="bobyqa", optCtrl=list(maxfun=2e5)))

# LR-test
pchisq(2*(logLik(full_model)[[1]]-logLic(reduced_model)[[1]]), 1, lower=F)
```

##### 2.3 Standard error of model parameters

Unfortunately `lme4` does not provides the analytical hessian matrix, we need to approximate the hessian matrix using a numerical method implemented in `numDeriv` on `R`. The following code provides the standard errors of standard deviations obtained from a `glmer` result:

```
library(numDeriv)
devfun = update(full_model, devFunOnly=T)
pars = getME(full_model, c("theta","fixef"))
hess = hessian(devfun, unlist(pars))
sdse = data.frame(sd=unlist(pars), se=sqrt(diag(solve(hess))))
```

The object `sdse` contains both standard deviations and their standard errors which were used to draw the forest plot.
