## Supplementary Note 2 for "The cellular immune response to COVID-19 deciphered by single cell multi-omics across three UK centres"

**Principal Investigators**
Stephen Baker, John Bradley, Gordon Dougan, Christoph Hess, Ian Goodfellow, Ravi Gupta, Nathalie Kingston, Paul J. Lehner, Paul A. Lyons, Nicholas J. Matheson, Willem H. Owehand, Caroline Saunders, Kenneth G.C. Smith, Charlotte Summers, James E. D. Thaventhiran, Mark Toshner and Michael P. Weekes

**CRF and Volunteer Research Nurses**
Ashlea Bucke, Jo Calder, Laura Canna, Jason Domingo, Anne Elmer, Stewart Fuller, Julie Harris, Sarah Hewitt, Jane Kennet, Sherly Jose, Jenny Kourampa, Anne Meadows, Criona O'Brien, Jane Price, Cherry Publico, Rebecca Rastall, Carla Ribeiro, Jane Rowlands, Valentina Ruffolo and Hugo Tordesillas

**Sample Logistics**
Ben Bullman, Benjamin J. Dunmore, Stuart Fawke, Stefan Gräf, Josh Hodgson, Christopher Huang, Kelvin Hunter, Emma Jones, Ekaterina Legchenko, Cecilia Matara, Jennifer Martin, Ciara O'Donnell, Linda Pointon, Nicole Pond, Joy Shih, Rachel Sutcliffe, Tobias Tilly, Carmen Treacy, Zhen Tong, Jennifer Wood and Marta Wylot

**Sample Processing**
Laura Bergamaschi, Ariana Betancourt, Georgie Bower, Aloka De Sa, Madeline Epping, Stuart Fawke, Oisin Huhn, Sarah Jackson, Isobel Jarvis, Jimmy Marsden, Francesca Nice, Georgina Okecha, Ommar Omarjee, Marianne Perera, Nathan Richoz, Rahul Sharma, and Lori Turner

**Clinical Data Collection**
Eckart M.D.D. De Bie, Katherine Bunclark, Masa Josipovic, Michael Mackay, Federica Mescia, Alice Michael, Sabrina Rossi, Mayurun Selvan, Sarah Spencer and Cissy Yong

**Royal Papworth Hospital ICU**
Ali Ansaripour, Alice Michael, Lucy Mwaura, Caroline Patterson and Gary Polwarth

**Addenbrooke's Hospital ICU**
Petra Polgarova and Giovanni di Stefano

**NIHR BioResource**
John Allison, Helen Butcher, Daniela Caputo, Debbie Clapham-Riley, Eleanor Dewhurst, Anita Furlong, Barbara Graves, Jennifer Gray, Tasmin Ivers, Mary Kasanicki, Emma Le Gresley, Rachel Linger, Sarah Meloy, Francesca Muldoon, Nigel Ovington, Sofia Papadia, Isabel Phelan, Hannah Stark, Kathleen E Stirrups, Paul Townsend, Neil Walker and Jennifer Webster

Stephen Baker[1, 2], John R. Bradley[2,8], Gordon Dougan[1, 2], Ian G Goodfellow[6], Ravindra K. Gupta[1, 2], Christoph Hess[1,2,15,16], Nathalie Kingston[7, 8], Paul J. Lehner[1, 2], Paul A. Lyons[1, 2], Nicholas J. Matheson[1, 2], Willem H. Owehand[7], Caroline Saunders[5], Kenneth G.C. Smith[1, 2], Charlotte Summers[,2,13,14,18], James E.D. Thaventhiran[1, 2, 11], Mark Toshner[2, 13, 14], Michael P. Weekes[1]

Ashlea Bucke[5], Jo Calder[5], Laura Canna[5], Jason Domingo[5], Anne Elmer[5], Stewart Fuller[5], Julie Harris[31], Sarah Hewitt[5], Jane Kennet[5], Sherly Jose[5], Jenny Kourampa[5], Anne Meadows[5], Criona O'Brien[31], Jane Price[5], Cherry Publico[5], Rebecca Rastall[5], Carla Ribeiro[5], Jane Rowlands[5], Valentina Ruffolo[5], Hugo Tordesillas[5],

Ben Bullman[1], Benjamin J. Dunmore[2], Stuart Fawke[20], Stefan Gräf[2,7,8], Josh Hodgson[2], Christopher Huang[2], Kelvin Hunter[1, 2], Emma Jones[19], Ekaterina Legchenko[2], Cecilia Matara[2], Jennifer Martin[2], Ciara O'Donnell[2], Linda Pointon[2], Nicole Pond[1, 2], Joy Shih[2], Rachel Sutcliffe[2], Tobias Tilly[2], Carmen Treacy[2], Zhen Tong[2], Jennifer Wood[2], Marta Wylot[26],

Laura Bergamaschi[1, 2], Ariana Betancourt[1, 2], Georgie Bower[1, 2], Aloka De Sa[2], Madeline Epping[1, 2], Stuart Fawke[20], Oisin Huhn[22], Sarah Jackson[2], Isobel Jarvis[2], Jimmy Marsden[2], Francesca Nice[29], Georgina Okecha[2], Ommar Omarjee[2], Marianne Perera[2], Nathan Richoz[2], Rahul Sharma[2], Lori Turner[1, 2]

Eckart M.D.D. De Bie[2], Katherine Bunclark[2], Masa Josipovic[30], Michael Mackay[2], Federica Mescia[1,2], Alice Michael[14], Sabrina Rossi[25], Mayurun Selvan[2], Sarah Spencer[4], Cissy Yong[25],

Ali Ansaripour[14], Alice Michael[14], Lucy Mwaura[14], Caroline Patterson[14], Gary Polwarth[14],

Petra Polgarova[18], Giovanni di Stefano[18],

John Allison[7,8], Helen Butcher[8,28], Daniela Caputo[8,28], Debbie Clapham-Riley[8,28], Eleanor Dewhurst[8,28], Anita Furlong[8,28], Barbara Graves[8,28], Jennifer Gray[8,28], Tasmin Ivers[8,28], Mary Kasanicki[8,18], Emma Le Gresley[8,28], Rachel Linger[8,28], Sarah Meloy[8,28], Francesca Muldoon[8,28], Nigel Ovington[7,8], Sofia Papadia[8,28], Isabel Phelan[8,28], Hannah Stark[8,28], Kathleen E Stirrups[7,8], Paul Townsend[7,8], Neil Walker[7,8], Jennifer Webster[8,28].

1 Cambridge Institute for Therapeutic Immunology and Infectious Disease, Jeffrey Cheah Biomedical Centre, University of Cambridge, Cambridge CB2 0AW, UK

2 Department of Medicine, University of Cambridge, Addenbrooke's Hospital, Cambridge CB2 0QQ, UK

3 MRC Biostatistics Unit, University of Cambridge, Cambridge Biomedical Campus, Cambridge CB2 0QQ, UK

4 Department of Clinical Biochemistry and Immunology, Addenbrooke's Hospital, Cambridge CB2 0QQ, UK

5 Cambridge Clinical Research Centre, NIHR Clinical Research Facility, Cambridge University Hospitals NHS Foundation Trust, Addenbrooke's Hospital, Cambridge CB2 0QQ, UK

6 Division of Virology, Department of Pathology, University of Cambridge, Addenbrooke's Hospital, Cambridge CB2 0QQ, UK

7 Department of Haematology, University of Cambridge, Cambridge Biomedical Campus, Cambridge CB2 0QQ, UK

8 NIHR BioResource, Cambridge University Hospitals NHS Foundation, Cambridge Biomedical Campus, Cambridge CB2 0QQ, UK.

9 Australian National Phenome Centre, Murdoch University, Murdoch, Western Australia WA 6150, Australia

10 Department of Clinical Biochemistry and Immunology, Addenbrooke's Hospital, Cambridge CB2 0QQ, UK

11 MRC Toxicology Unit, School of Biological Sciences, University of Cambridge, Cambridge CB2 1QR, UK

12 R&D Department, Hycult Biotech, 5405 PD Uden, The Netherlands

13 Heart and Lung Research Institute, Cambridge Biomedical Campus, Cambridge CB2 0QQ, UK

14 Royal Papworth Hospital NHS Foundation Trust, Cambridge Biomedical Campus, Cambridge CB2 0QQ, UK

15 Department of Biomedicine, University and University Hospital Basel, 4031Basel, Switzerland

16 Botnar Research Centre for Child Health (BRCCH) University Basel & ETH Zurich, 4058 Basel, Switzerland

17 These authors contributed equally

18 Addenbrooke's Hospital, Cambridge CB2 0QQ, UK

19 Department of Veterinary Medicine, Madingley Road, Cambridge, CB3 0ES, UK

20 Cambridge Institute for Medical Research, Cambridge Biomedical Campus, Cambridge CB2 0XY, UK

21 Cancer Research UK, Cambridge Institute, University of Cambridge CB2 0RE, UK

22 Department of Obstetrics & Gynaecology, The Rosie Maternity Hospital, Robinson Way, Cambridge CB2 0SW, UK

23 Centre for Molecular Medicine and Innovative Therapeutics, Health Futures Institute, Murdoch University, Perth, WA, Australia

24 Cambridge and Peterborough Foundation Trust, Fulbourn Hospital, Fulbourn, Cambridge CB21 5EF, UK

25 Department of Surgery, Addenbrooke's Hospital, Cambridge CB2 0QQ, UK

26 Department of Biochemistry, University of Cambridge, Cambridge, CB2 1QW, UK

27 Centre of Computational and Systems Medicine, Health Futures Institute, Murdoch University, Harry Perkins Building, Perth, WA 6150, Australia

28 Department of Public Health and Primary Care, School of Clinical Medicine, University of Cambridge, Cambridge Biomedical Campus, Cambridge, UK

29 Cancer Molecular Diagnostics Laboratory, Department of Oncology, University of Cambridge, Cambridge CB2 0AH, UK

30 Metabolic Research Laboratories, Wellcome Trust-Medical Research Council Institute of Metabolic Science, University of Cambridge, Cambridge CB2 0QQ, UK

31 Department of Paediatrics, University of Cambridge, Cambridge Biomedical Campus, Cambridge, CB2 0QQ, UK
